# Comprehensive baseline ctDNA characterization in two biomarker-selected Phase 1/2 studies using genomic and methylation profiling

**DOI:** 10.1101/2024.11.11.24316049

**Authors:** Ian M. Silverman, Joseph D. Schonhoft, Benjamin Herzberg, Arielle Yablonovitch, Errin Lagow, Sunantha Sethuraman, Danielle Ulanet, Julia Yang, Insil Kim, Paul Basciano, Michael Cecchini, Elizabeth Lee, Stephanie Lheureux, Elisa Fontana, Benedito A. Carneiro, Jorge S. Reis-Filho, Timothy A. Yap, Michael Zinda, Ezra Y. Rosen, Victoria Rimkunas

## Abstract

The development of DNA damage response (DDR)-directed therapies is a major area of clinical investigation, yet to date Poly (ADP-ribose) polymerase (PARP) inhibitors remain the only approved therapy in this space. Major challenges to DDR-targeted therapies in the post-PARPi era are the context dependency of DDR alterations and the presence of pre-existing resistance in this heavily pre-treated population. To that end, we used a contemporary platform to analyze pre-treatment circulating tumor DNA (ctDNA) samples from 173 patients enrolled onto two Phase 1/2 trials harboring pathogenic variants (PVs) in DDR genes. Baseline ctDNA analysis revealed a wealth of insights, including circulating tumor fraction estimation, impact of clonal hematopoiesis, PV allelic status, homologous recombination deficiency (HRD) signatures and presence of pre-existing resistance. HRD reversions were detected in 44% of evaluable patients and included large genomic rearrangements leading to deletion of whole or partial exons. We also discovered reversion of *ATM* in two patients previously treated with platinum chemotherapy, which has not previously been described. This study showcases the genomic complexity of DDR-altered tumors, revealed through baseline ctDNA profiling, an understanding of which is crucial for the future clinical development of novel DDR-directed therapies and combinations.

## INTRODUCTION

Homologous recombination deficiency (HRD) is commonly observed in BRCA-associated tumors (ovarian, breast, prostate and pancreatic) typically harboring biallelic loss-of-function (LoF) of HRD-associated genes; *BRCA1*, *BRCA2*, *PALB2,* and the *RAD51* paralogs (*RAD51B/C/D*, collectively *RAD51*p). HRD tumors are exquisitely sensitive to PARPi/platinum therapy, but therapeutic resistance invariably develops. Multiple mechanisms of resistance have been described, including reversion of the PV, Shieldin complex loss, increased drug efflux, lineage plasticity, and mutations in *PARP1*, among others^1–3^.

Among these mechanisms, reversion mutations, which restore HR activity, are the most well-characterized source of resistance to PARPi/platinum therapies^4–10^. Reversions were initially identified in high-grade ovarian tumors treated with platinum-based chemotherapy^6,7,11,12^, and subsequent studies have documented reversions across breast, pancreatic, and prostate tumors^4,5,8,13^. More than one thousand reversions, primarily in *BRCA1/2*, and to a lesser extent in *PALB2* and *RAD51p,* have been detected in tissue and plasma samples collected from patients with BRCA-associated tumors treated previously with PARPi/platinum therapies^4–10^. The presence of reversions consistently correlates with poor response and survival^4,5,8,13–15^. Conversely, castrate-resistant prostate tumors harboring unrevertible PVs (e.g. homozygous deletions in *BRCA2*) typically respond exceptionally well to PARPi with prolonged disease control^16^.

Most previously characterized reversions are short variants that restore the coding sequences with minimal changes. However, larger, more complex genomic rearrangements significantly altering the amino acid sequence and domain structures in *BRCA1/2* have also been reported^4,5,13,17^. The specific functional and clinical consequences of these complex reversions, notably the degree of HR proficiency, and how these sequelae may vary by reversion type and mechanism, have not been characterized to date.

Gaining a better understanding of the spectrum and development of reversion mutations is critical for future therapeutic strategies to address PARPi/platinum therapy resistance. Clinical trials are evaluating new agents in genomically defined subsets of patients with BRCA-associated tumors previously treated with PARPi/platinum therapy, yet few studies characterize HR status or reversion mutations at the time of enrollment. Without an understanding of a patient’s contemporaneous HR status, clinical trial efficacy results will be misinterpreted, and useful therapies underdeveloped due to a perceived lack of activity. A deeper understanding of the functional consequences of reversions may lead to more personalized therapies, including rechallenge with alternative or next generation DNA damage response (DDR) targeted agents in the appropriate clinical setting.

Obtaining temporally matched tissue biospecimens to accurately assess HR status in these patients with advanced cancers has been challenging. Next-generation sequencing (NGS) of post-progression biopsies is rarely performed, and when it is, sampling biases can limit the detection of heterogeneous, polyclonal resistance. Conversely, circulating tumor DNA (ctDNA) assays can detect diverse resistance mutations without sampling biases. Historically, ctDNA assays have been narrowly focused on exons and consequently lacked the ability to detect most large genomic rearrangements (LGR), and there is a high risk of false negative results for patients with low ctDNA burden^18^. Therefore, in addition to resistance mechanisms, interpretation of the results in the context of other covariates such as circulating tumor fraction (TF) and HRD signatures may be necessary. Due to a lack of sufficient methods and implementation, historical analyses often fail to accurately reflect the true frequency of reversions and the nature of acquired resistance in the post-PARPi/platinum setting.

Therefore, we conducted a comprehensive analysis of ctDNA samples collected from patients enrolled on two Phase 1/2 clinical trials evaluating the Ataxia Telangiectasia and RAD3-related kinase (ATR) inhibitor, camonsertib. We used this large cohort of molecularly selected patients consisting of BRCA-associated and other tumor types to evaluate the performance of a contemporary genomic and epigenomic ctDNA assay with enhanced gene coverage (including introns) for detecting reversions, HRD signatures, PV allele status, presence of CH, and differentially methylated regions for accurate quantitation of TF.

## RESULTS

### Cohort description and baseline genomic analysis

Blood samples were obtained from patients participating in the Phase 1/2 clinical trials TRESR (NCT04497116) and ATTACC (NCT04972110) evaluating the ATR inhibitor (ATRi), camonsertib as a monotherapy, in combination with gemcitabine (TRESR), or in combination with one of three approved PARPi - talazoparib (TRESR) or niraparib, olaparib (ATTACC). A key molecular eligibility criterion for these trials was the presence of a PV in an HR or DDR gene predicted to be synthetic lethal with ATRi or ATRi/PARPi as documented by a local NGS test (see methods; **Fig. 1a**). Patients with any solid tumor were eligible for these trials, however the cohort was enriched for patients with BRCA-associated tumor types. Consequently, most patients were previously treated with PARPi, platinum, or both classes of agents (**Table 1**). We analyzed ctDNA in baseline plasma samples from 173 patients using a commercially available assay (Guardant Infinity^TM^) that includes 753 genes, covers all introns (excluding repetitive sequences) and exons of *BRCA1/2*, and estimates tumor fraction (TF) from differentially methylated tumor-derived DNA. We also analyzed variants from baseline buffy coats from 155 patients using the same platform to adjudicate variants derived from clonal hematopoiesis (CH; **Fig. 1b**).

**Fig. 1:**
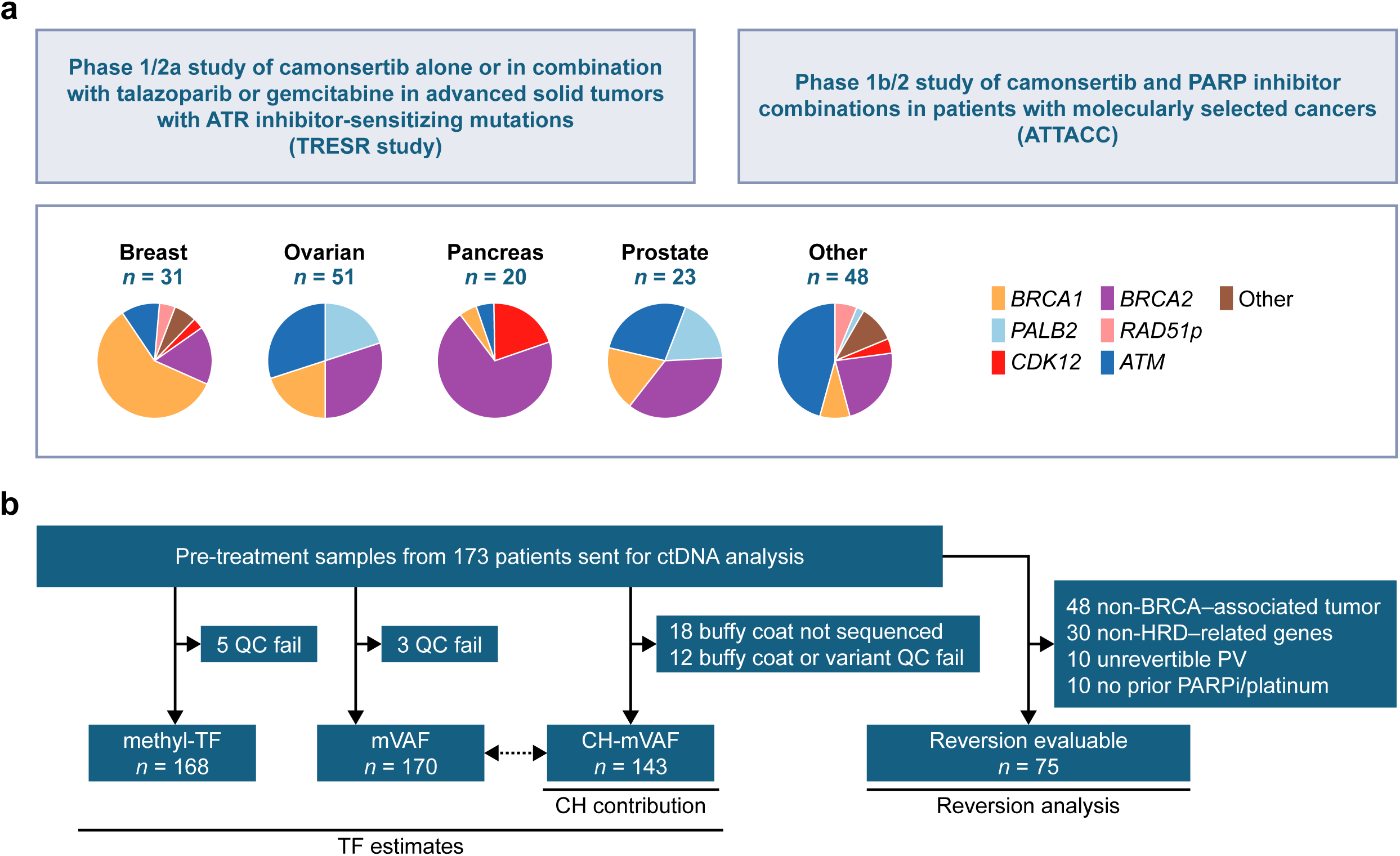
Study overview. **a)** Overview of patients included in the study enrolled on TRESR and ATTACC and the breakdown of enrollment genotypes in the most prevalent tumor types. **b)** Consort diagram of evaluable patients for the different analysis subsets including tumor fraction estimates, clonal hematopoiesis evaluation and reversion analysis. BRCA-associated tumors refer to breast, ovarian, prostate, and pancreatic cancers. RAD51p = RAD51 paralogs; includes RAD51B, RAD51C and RAD51D. ATR, ataxia telangiectasia and RAD3-related kinase; CH, clonal hematopoiesis; ctDNA, circulating tumor DNA; HRD, homologous recombination deficiency; methyl-TF, methylation-based TF; mVAF, mean variant allele frequency; PARPi, Poly (ADP-ribose) polymerase (PARP) inhibitors inhibitor; PV, pathogenic variant; QC, quality control; TF, tumor fraction.

**Table 1:**
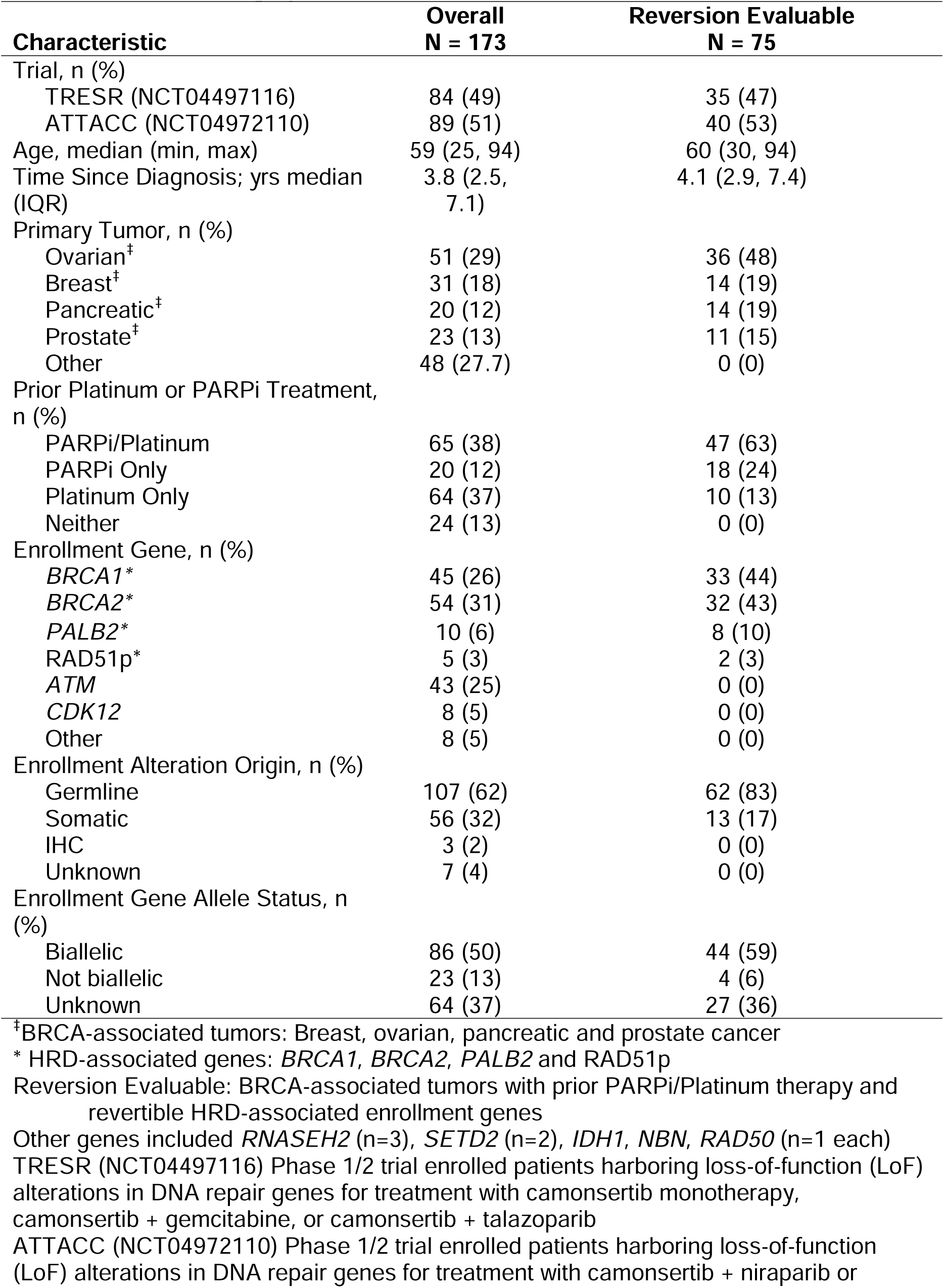
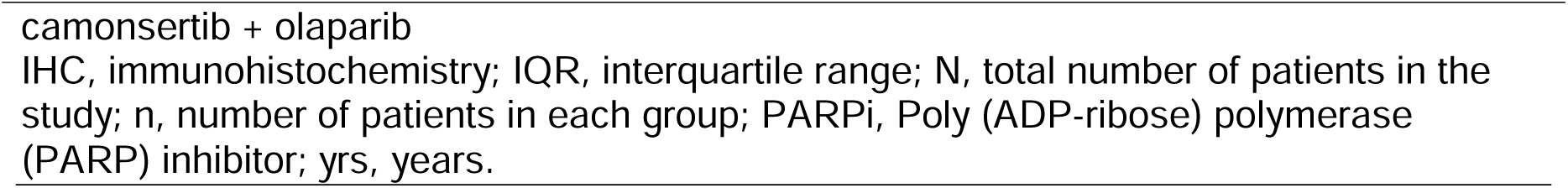
Patient demographics.

| Characteristic | Overall<br>N = 173 | Reversion Evaluable<br>N = 75 |
| --- | --- | --- |
| Trial, n (%) |  |  |
| TRESR (NCT04497116) | 84 (49) | 35 (47) |
| ATTACC (NCT04972110) | 89 (51) | 40 (53) |
| Age, median (min, max) | 59 (25, 94) | 60 (30, 94) |
| Time Since Diagnosis; yrs median (IQR) | 3.8 (2.5, 7.1) | 4.1 (2.9, 7.4) |
| Primary Tumor, n (%) |  |  |
| Ovarian<sup>‡</sup> | 51 (29) | 36 (48) |
| Breast<sup>‡</sup> | 31 (18) | 14 (19) |
| Pancreatic<sup>‡</sup> | 20 (12) | 14 (19) |
| Prostate<sup>‡</sup> | 23 (13) | 11 (15) |
| Other | 48 (27.7) | 0 (0) |
| Prior Platinum or PARPi Treatment, n (%) |  |  |
| PARPi/Platinum | 65 (38) | 47 (63) |
| PARPi Only | 20 (12) | 18 (24) |
| Platinum Only | 64 (37) | 10 (13) |
| Neither | 24 (13) | 0 (0) |
| Enrollment Gene, n (%) |  |  |
| *BRCA1*<sup>*</sup> | 45 (26) | 33 (44) |
| *BRCA2*<sup>*</sup> | 54 (31) | 32 (43) |
| *PALB2*<sup>*</sup> | 10 (6) | 8 (10) |
| *RAD51p*<sup>*</sup> | 5 (3) | 2 (3) |
| *ATM* | 43 (25) | 0 (0) |
| *CDK12* | 8 (5) | 0 (0) |
| Other | 8 (5) | 0 (0) |
| Enrollment Alteration Origin, n (%) |  |  |
| Germline | 107 (62) | 62 (83) |
| Somatic | 56 (32) | 13 (17) |
| IHC | 3 (2) | 0 (0) |
| Unknown | 7 (4) | 0 (0) |
| Enrollment Gene Allele Status, n (%) |  |  |
| Biallelic | 86 (50) | 44 (59) |
| Not biallelic | 23 (13) | 4 (6) |
| Unknown | 64 (37) | 27 (36) |

<sup>‡</sup>BRCA-associated tumors: Breast, ovarian, pancreatic and prostate cancer
<sup>\*</sup> HRD-associated genes: *BRCA1*, *BRCA2*, *PALB2* and *RAD51p*
Reversion Evaluable: BRCA-associated tumors with prior PARPi/Platinum therapy and revertible HRD-associated enrollment genes
Other genes included *RNASEH2* (n=3), *SETD2* (n=2), *IDH1*, *NBN*, *RAD50* (n=1 each) TRESR (NCT04497116) Phase 1/2 trial enrolled patients harboring loss-of-function (LoF) alterations in DNA repair genes for treatment with camonsertib monotherapy, camonsertib + gemcitabine, or camonsertib + talazoparib ATTACC (NCT04972110) Phase 1/2 trial enrolled patients harboring loss-of-function (LoF) alterations in DNA repair genes for treatment with camonsertib + niraparib or
camonsertib + olaparib
IHC, immunohistochemistry; IQR, interquartile range; N, total number of patients in the study; n, number of patients in each group; PARPi, Poly (ADP-ribose) polymerase (PARP) inhibitor; yrs, years.

### Baseline ctDNA tumor fraction and correlation with disease burden

ctDNA was detected by methylation-based analysis in 94% (158/168; 5 failed methylation QC) of baseline plasma samples with a median methylation-based tumor fraction (methyl-TF) of 6.2% (IQR: 24.7%; **Fig. 2a**). methyl-TF varied across tumor types, with the highest observed in prostate cancer (median methyl-TF 30.9%; IQR 65.0%) and lowest in ovarian cancer (median methyl-TF 3.6% IQR 9.28%). In contrast to methyl-TF, the variant-based approach detected ctDNA in fewer patients, with only 68% (115/170; 3 failed variant QC) of baseline plasma samples or 66% (95/143) after filtering additional variants derived from CH using paired buffy-coat sequencing (**Supplementary Fig. 1a, B**). Mean variant allele frequencies (mVAF) were similar between plasma-only (median mVAF 6.4%: IQR 13.3%) and CH-filtered (median CH-mVAF 6.6%: IQR 14.2%) methods. For samples with detectable ctDNA by both methylation- and variant-based methods, mVAF and CH-mVAF showed strong correlation with methyl-TF, with CH-mVAF showing the stronger correlation (R=0.84; p < 2.2e-16, Pearson’s correlation coefficient) **(Supplementary Fig. 1c, d)**. mVAF and CH-mVAF were highly correlated (r=0.98; p<2.2e-16, Pearson’s correlation coefficient), with modest differences in 15 samples (**Supplementary Fig. 1e**). However, in 5 patients, variants were no longer monitorable after CH-filtering (**Supplementary Fig. 1f**). A modest correlation was observed between baseline TF and target lesion sum (RECIST V1.1) but this analysis was confounded by the absence of quantification of non-target lesions (**Fig. 2b and Supplementary Fig. 2a, b**). No relationships between baseline TF and CA-125 (ovarian only) or PSA (prostate only) were observed (**Supplementary Fig. 2c**).

**Fig. 2:**
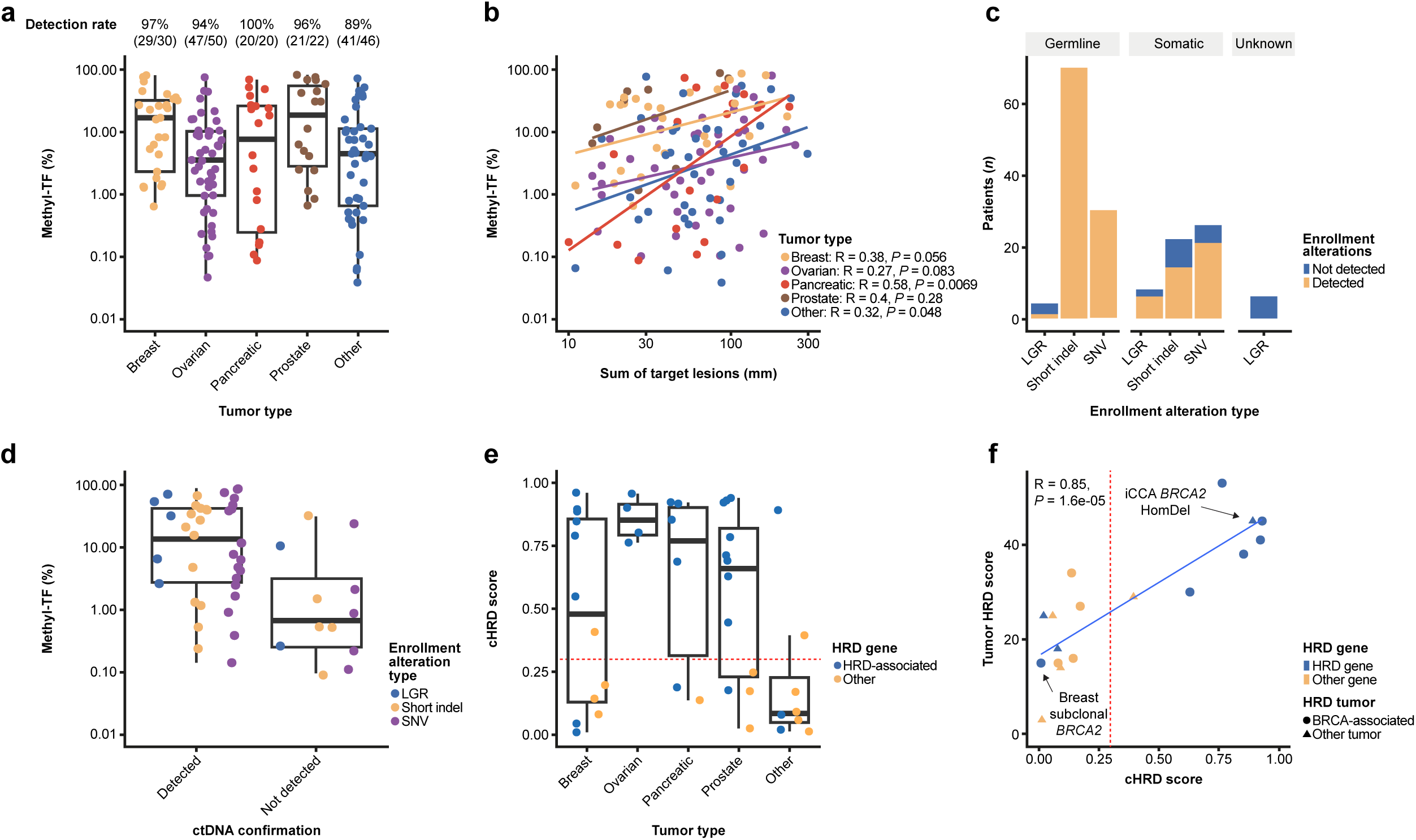
Baseline characterization of plasma samples. **a)** methyl-TF detection and estimation by tumor type. **b)** Correlation between methyl-TF and baseline sum of target lesion diameters by tumor type. **c)** Detection of the enrollment PV by alteration class (LGR [deletions and structural alterations], short indels and SNVs] and alteration origin. **d)** methyl-TF in patients with detected vs. not detected somatic enrollment PVs. **e)** cHRD scores by tumor type and enrollment gene category for samples with >25% methyl-TF. Horizontal read line is the cHRD positivity threshold. **f)** Correlation between tumor HRD and cHRD scores in evaluable samples. Horizontal read line is the cHRD positivity threshold. HRD-associated genes, *BRCA1, BRCA2, PALB2 and* RAD51p; BRCA-associated tumor, breast, ovarian, pancreatic and prostate cancers. cHRD, ctDNA-based HRD; ctDNA, circulating tumor DNA; HomDel, homozygous deletion; HRD, homologous recombination deficiency; iCCA, intrahepatic cholangiocarcinoma; indel, insertion or deletion; LGR, large genomic rearrangements; methyl-TF, methylation-based TF; PV, pathogenic variant; SNVs, Single nucleotide variants; TF, tumor fraction; vs., versus.

### Detection of enrollment PV, allelic status and HR status in ctDNA

The enrollment PV (i.e. the BRCA1/2 or other DDR gene alteration for which patient entry into the trial population was based) was confirmed in baseline ctDNA from 142/166 evaluable patients (86%, 3 excluded for immunohistochemistry [IHC] enrollment, 4 excluded for QC failures; **Fig. 2c**). Baseline methyl-TF was significantly higher in samples where the somatic enrollment PV was detected versus (vs.) undetected (median methyl-TF 15.1% vs. 0.6%; p = 0.0009, Wilcoxon rank sum test; **Fig. 2d**). In 3 cases with undetected somatic enrollment PVs but high methyl-TF, enrollment PVs were also undetected in tissue NGS, corroborating the ctDNA findings. LGRs, including structural variants and deletions were confirmed for 39% (7/18) of patients. Excluding germline PVs, baseline methyl-TF was also significantly higher in those with detected vs. undetected enrollment LGR PVs (41.4% vs 0.03%, p=0.04, Wilcoxon rank sum test; **Supplementary Fig. 3a**). Collectively, these data suggest that somatic PVs may be missed due to low TF or due to heterogeneous local tests and subclonal alterations in cases with high TF.

Biallelic LoF of DDR genes is thought to be a molecular determinant of response to synthetic lethal targeted therapies such as PARPi^19^, although measurement of this complex biomarker is typically restricted to tumor analyses and is often inferred by the presence of a germline mutation in a BRCA-associated tumor type, rather than directly observed. We investigated whether biallelic LoF could be resolved in ctDNA. Biallelic LoF was detected in 30% (43/142) of baseline samples with a detected enrollment PV, comprising i) PV accompanied by loss of heterozygosity (LOH; n=24), ii) compound heterozygous PVs (n=16), or iii) homozygous deletions (n=3; **Supplementary Fig. 3b**). Consistent with detection of PVs, methyl-TF was significantly higher in patients with biallelic LoF detected vs. not detected (37.5% vs. 4.24%; p=3.1e-10 Wilcoxon rank sum test; **Supplementary Fig. 3c**). When comparing ctDNA-based detection to allelic status derived from tissue NGS (methods), biallelic LoF was confirmed in 44% (33/75) of cases known to have biallelic LoF, and only 6% (1/16) of cases known to have non-biallelic LoF (**Supplementary Fig. 3d**). In the one discordant case, tissue NGS detected a germline *BRCA2* PV and *BRCA2* LOH, however, the allele containing the *BRCA2* PV was deleted in tumor cells and therefore classified as non-biallelic. Importantly, ctDNA analysis identified 9 patients as having biallelic LoF who were indeterminate by tissue NGS due to lack of sufficient sample quantity or quality, indicating potential clinical utility when tumor-based methods are unavailable.

Genomic signatures of HRD are another complex biomarker typically restricted to tumor tissue analysis, and are predictive of response to PARPi^20^. We assessed feasibility of detecting genomic signatures of HRD utilizing ctDNA. A validated probabilistic model (see methods) was utilized. In cases with a methyl-TF >25% threshold, ctDNA-based HRD (cHRD) positivity (≥ 0.3) was detected in baseline plasma samples from 60% (25/42) of all patients and 85% (22/26) of patients with BRCA-associated tumors harboring HRD-associated gene alterations (*i.e. BRCA1, BRCA2, PALB2 and RAD51p*). (**Fig. 2e**). Amongst the non-BRCA-associated tumor types, samples from 2 patients were cHRD positive, including an intrahepatic cholangiocarcinoma patient with a *BRCA2* homozygous deletion and a cervical carcinoma patient with a biallelic *ATM* somatic mutation. Further restricting the analysis to the BRCA-associated tumors harboring HRD-associated gene alterations with confirmed biallelic LoF in tissue or ctDNA, the cHRD positivity rate increased to 93% (14/15) and 94% (17/18), respectively (**Supplementary Fig. 4a, b**). Finally, in the subset of patients with available tumor HRD scores (n=17; see Methods), the HRD scores between ctDNA and tumor were significantly correlated (R=0.85, p=1.6e-5; Pearson’s correlation coefficient; **Fig. 2f**).

### Reversion detection in patients with HRD associated tumors and genes

To characterize reversion alterations and define the reversion evaluable cohort (n=75), we limited the analysis to plasma samples (including 104 available on-treatment timepoints) collected from patients with BRCA-associated tumors and HRD-associated gene alterations previously treated with PARPi/platinum therapy while eliminating cases with PVs that cannot be reverted (i.e., homozygous deletions and rearrangements; **Fig. 1b**). As expected, no reversions were found in the unrevertible (n=10) or PARPi/platinum-naive (n=10) cases. Reversions were identified in 44% (33/75) within the reversion evaluable cohort, including 68 unique reversion alterations detected in at least one sample from each patient (**Fig. 3a and Supplementary Table 1).** In 39% (13/33) of patients, the reversions were polyclonal in nature with the number of unique reversions per patient ranging from 1–10, with the most extreme examples being 2 prostate cancer patients with 8 and 10 unique reversions. The cancer types with the highest rate of reversions were breast (64%, 9/14) and prostate (64%, 7/11), followed by pancreatic (43%, 6/14) and ovarian cancer (31%, 11/36; **Fig. 3b**). Reversions were found regardless of prior therapy, with PARPi associating more strongly: PARPi only (61%; 11/18), PARPi/platinum (40%, 19/47) and platinum only (30%; 3/10; **Supplementary Fig. 5a, b**). Reversions were identified across *BRCA2* (62%, 20/32), *BRCA1* (27%, 9/33), and *PALB2* (50%, 4/8) but absent in the two patients harboring *RAD51*p PVs (**Fig. 3c**). The frequency of reversions did not differ between PVs of germline (45%; 28/62) or somatic (37%; 5/13) origin. However, there was an enrichment of reversions if the enrolling PV identified in tissue was biallelic (50%, 22/44) when compared to the non-biallelic group (0%, 0/4; **Supplementary Fig. 5c**). This observation held true for the cohort of biallelic patients identified by ctDNA with reversions detected in 68% (15/22) of patients with ctDNA-based biallelic status vs. 34% (18/53) of those with biallelic status was not detected (**Supplementary Fig. 5d**). Furthermore, reversions were detected in 50% (10/20) of samples predicted to be HRD positive underlining the importance of reversion detection for contemporaneous HR status determination (**Supplementary Fig. 5e**).

**Fig. 3:**
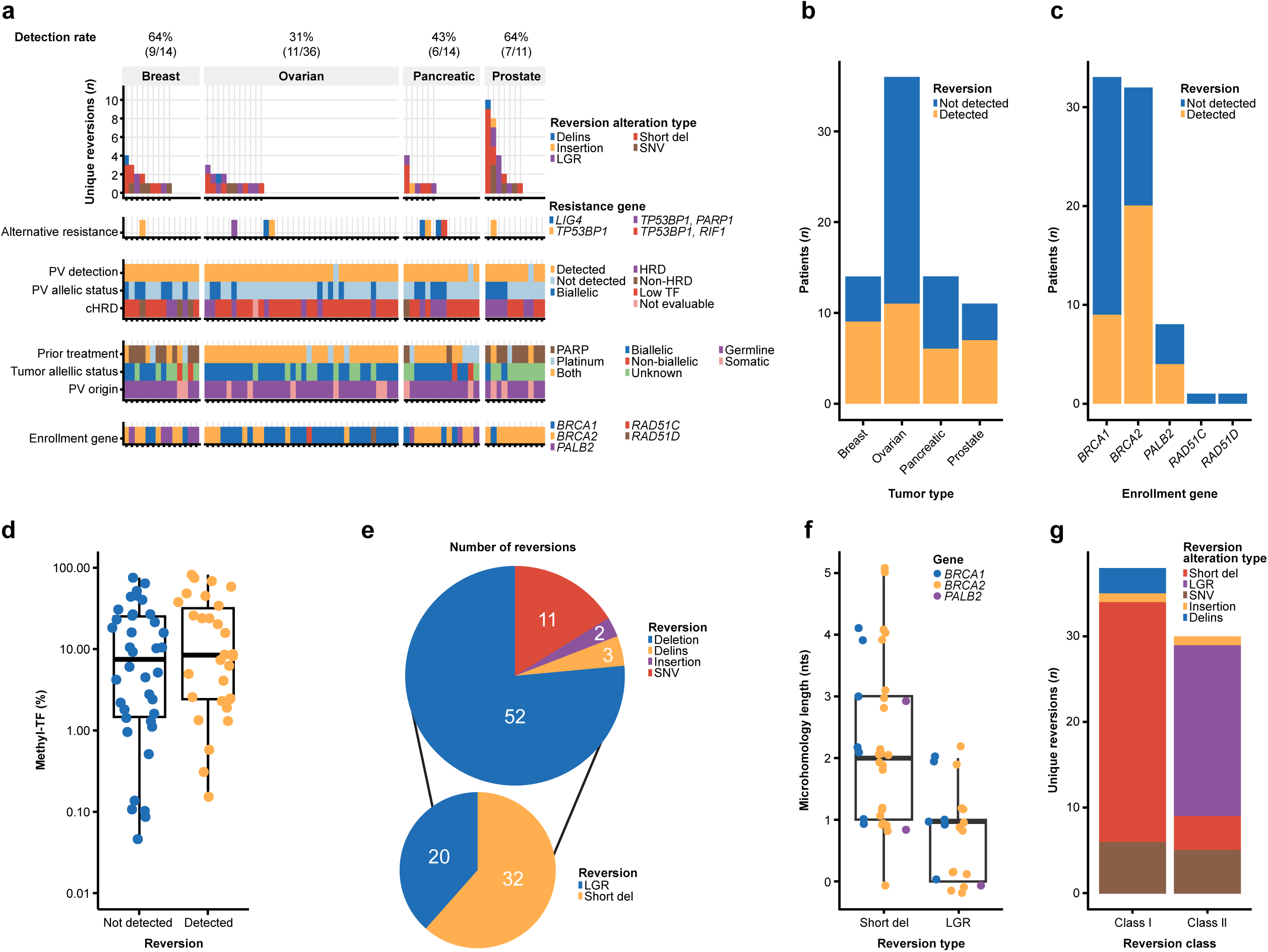
Reversion detection in patients with BRCA-associated tumors and HRD PVs. **a)** Unique reversion count in the 75 reversion evaluable patients. **b)** Reversion detection by tumor type and **c)** genotype. **d)** methyl-TF in patients with and without detected reversions. **e)** Reversion alteration classification and subclassification of deletions with varying lengths. **f)** Microhomology length in patients with short deletions (>1nt) vs. LGR deletions. **g)** Reversion alteration classification for Class I and Class II reversions. cHRD, ctDNA-based HRD; ctDNA, circulating tumor DNA; del, deletions; delins, deletion-insertions; HRD, homologous recombination deficiency; LGR, large genomic rearrangements; methyl-TF, methylation-based TF; NA, not applicable; nt, nucleotide; PARPi, Poly (ADP-ribose) polymerase (PARP) inhibitors; PV, pathogenic variant; SNVs, Single nucleotide variants; TF, tumor fraction; vs., versus.

Importantly, the median methyl-TF was not significantly different in patients with detected reversions (8.6%) compared with those with no reversions detected (7.7%: p=0.27, Wilcoxon rank sum test; **Fig. 3d**). However, no reversions were detected in 9 patients with undetected or extremely low tumor shedding (methyl-TF < 0.15%) suggesting a lower limit of TF at which reversions can be detected.

Deletions were the most common reversion alteration type, accounting for 76% (52/68) of all reversions and comprising 32 short deletions and 20 LGRs (**Fig. 3e)**. Single nucleotide variants (SNVs; n=11), short insertions (n=2) and deletion-insertions (delins; n=3) were also observed. LGR reversions had a median size of 2,534 nucleotides (nts; range: 99–15,828 nt) and were detected in 15 patients including those enrolled with PVs in *BRCA1* (56%, 5/9), *BRCA2* (47% 9/19) and *PALB2* (25% 1/4; **Fig. 3a and Supplementary Fig. 5f**). In 9 patients, LGRs were the only class of reversions, 6 of which enrolled with *BRCA2* PVs.

In HR deficient tumors, microhomology-mediated end joining (MMEJ) is active and results in error prone DNA repair leading to short deletions with microhomology, which has been reported to be a common characteristic of reversions^5,21^. We evaluated whether there was evidence of MMEJ through the presence of short homologous sequences at breakpoints. When defined as at least 1nt overlap, microhomology was observed in 97% (31/32) of short deletions and 60% (12/20) of LGRs (**Fig. 3f**). Furthermore, microhomology length was significantly longer in short deletions vs. LGRs (median 2 nts vs. 1 nt, p=6.5e-5, Fisher’s exact test) suggesting potential alternative mechanisms for biogenesis of LGR reversions.

### Reversion classification by impact on coding sequence

Within our dataset, we observed two distinct classes of reversions, differentiated by the extent of their impact on protein sequence (**Fig. 3g**). Class I (n=38) reversions were mostly short deletions and SNVs leading to minimal changes in protein sequence and were representative of most known reversions, due to the ease of detection with exon targeted NGS panels (**Supplementary Table 1**). Given the minimal changes to the protein sequence, these reversions are unlikely to have a major impact on protein function.

In contrast, Class II (n=30) reversions comprised mostly LGRs and result in genomic or post-transcriptional deletion of whole or partial exons^13^ (**Fig. 4a, c**). These complex reversions were detected in 58% (19/33) of patients with reversions, and of those, 74% (14/19) had exclusively Class II (**Fig. 3a**). Class II reversions were observed across BRCA-associated tumors and HRD-associated genes with the highest prevalence in *BRCA2* altered tumors (38%; 12/32) and in ovarian cancers (22%; 8/36). Given the technical challenge of identifying these alterations, they are less well described in the literature and the degree of their impact on restoration of HR function may vary depending on the location and extent of protein sequence change.

**Fig. 4:**
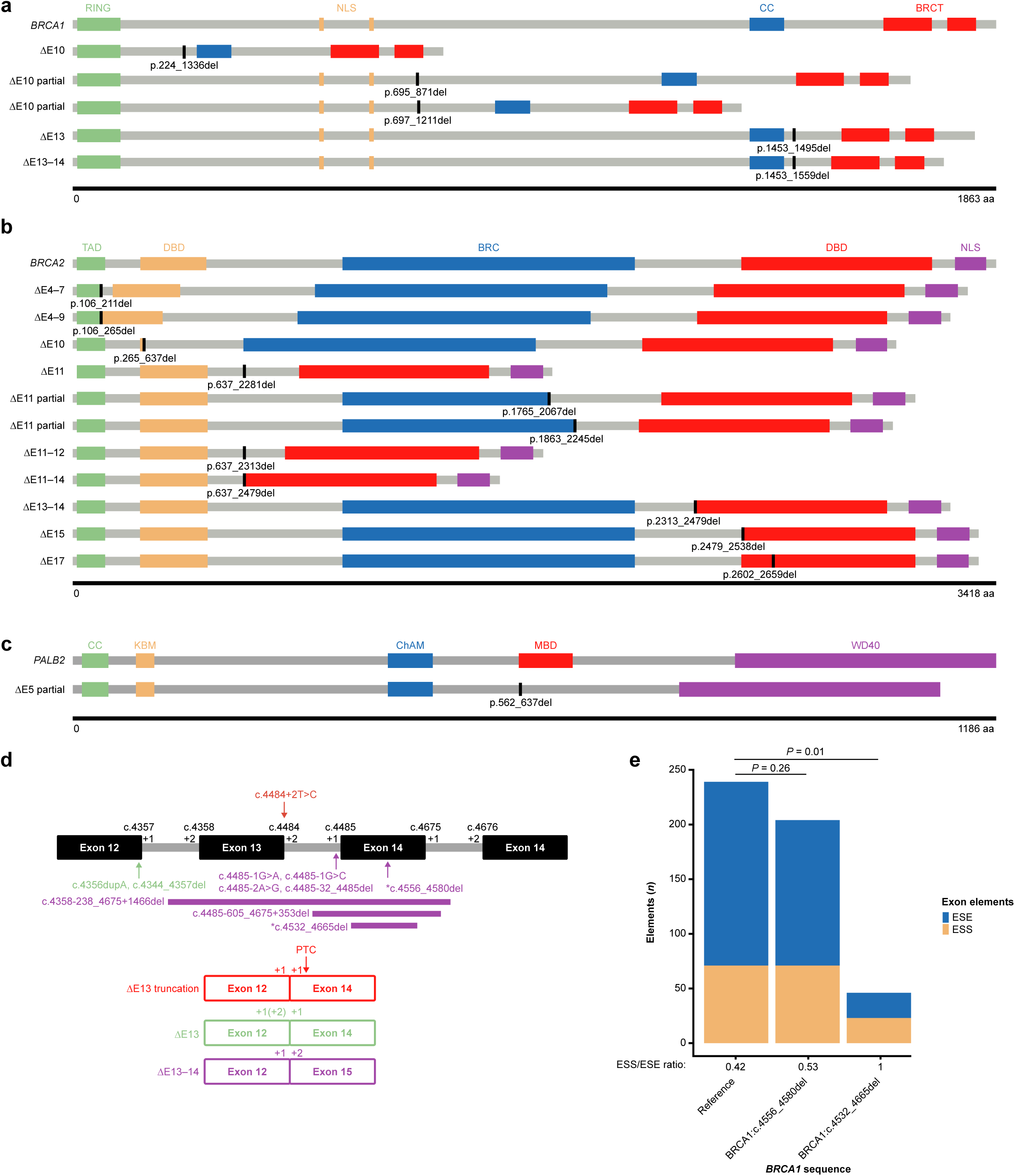
Class II reversions impact on protein structure. **a, b, c)** Schematic of the predicted protein structure for class II reversions in **a)** BRCA1, **b)** BRCA2, and **c)** PALB2. **d)** Schematic of the patient with *BRCA1* PV (c.4484+2T>C) and the 10 secondary variants, 8 of which were classified as Class II reversions. Red indicates the PV and predicted ΔE13 truncation; green indicates the ΔE13 reversions and purple represents the ΔE13-14 reversion. **e)** EX-SKIP analysis of the two secondary variants not-predicted to be reversions demonstrated specific loss of ESE in Exon 14 and predicted to have in-frame splicing leading to ΔE13-14. BRC, BRC^Δ^repeats; BRCT, BRCA1 c-terminal; CC, coiled coil; ChAM, chromatin binding motif; DBD, DNA binding domain; del, deletions; ESE, exon splicing enhancer; ESS, exon splicing silencers; KBM, KEAP1 binding motif; MBD, MRG15 binding domain; NLS, nuclear localization signal; PTC, premature termination codon; PV, pathogenic variant; RING, ring finger; TAD, transcriptional activation domain; WD40, WD40 repeats.

The complexity of reversions can be highlighted by specific cases. A patient with castration-resistant prostate cancer, previously treated with PARPi/platinum, harbored a somatic biallelic *BRCA1* PV (c.4484+2T>C) predicted to lead to splicing of exon 12 to 14 (ΔE13 truncation) resulting in a premature stop codon (**Fig. 4d**). ctDNA analysis revealed 10 additional *BRCA1* alterations including 3 SNVs, 3 short deletions, 1 insertion and 3 LGRs, 8 of which were predicted to be Class II reversions. Among the reversions, 6 were predicted to delete or disrupt the exon 14, 3’ splice site yielding in-frame splicing of exon 12 to exon 15 (ΔE13-14), and two short deletions proximal to the 5’ splice site of exon 12 were predicted to alter the reading-frame and yield in-frame splicing to exon 14 (ΔE13). We observed 2 additional deletions (25 and 134 nt, respectively) within exon 14 for which a reversion mechanism could not clearly be ascribed. We hypothesized that these mutations result in loss of an exon splicing enhancer (ESE) within exon 14 resulting in skipping of exons 13 and 14 leading to in-frame splicing of exons 12 to 15 (ΔE13-14). Sequence analysis revealed a specific loss of ESE elements relative to exon splicing silencers (ESS), supporting this putative mechanism (**Fig. 4e**). Collectively, the variety of Class II reversions identified in ctDNA from this patient highlight the complexity of reversion detection and the need for fit-for-purpose diagnostics and sophisticated algorithms for reversion detection and classification.

### Alternative resistance to platinum and PARPi therapy

In addition to reversion mutations in *BRCA1/2*, loss of function alterations in *TP53BP1*, *SHLD1/2*, *PARP1*, *RIF1*, *LIG4* and *XRCC4* are hypothesized to cause resistance to PARPi/platinum therapies^13,22–25^. Among the 75 reversion evaluable patients, alterations in putative resistance genes were detected at baseline in 9 patients (**Fig. 3a**), while zero were detected in samples from the 10 patients with unrevertible PV alterations or 10 platinum/PARPi naïve patients. *TP53BP1* was altered in 6 patients (including 4 patients with *BRCA1* PVs), followed by *LIG4* in 3 patients and *RIF1* and *PARP1* in 1 patient each. All patients had evidence for biallelic *BRCA1* or *BRCA2* LoF by either tumor NGS (n=8) or ctDNA (n=6), and 6 were cHRD positive. In 55% (5/9) of patients with alternative resistance mutations, reversions were also detected. In a specific case, a patient with g*BRCA1* pancreatic cancer did not exhibit detectable reversions, but had 22 unique *TP53BP1* mutations, *TP53BP1* LOH, and one *RIF1* mutation, none of which were found in matched buffy coats (**Supplemental Fig. 6**). This patient had undergone treatment with FOLFIRINOX for 3.7 months (best response of stable disease), olaparib for 2.5 months (discontinued due to progressive disease), and FOLFOX for 1.4 months (stopped due to toxicity) in the 8 months leading up to sample collection.

### Detection of novel reversions in two patients with somatic ATM alterations

Prior to this study, reversions were reported in only select HRD-associated genes (e.g. *BRCA1/2*, *PALB2*, *RAD51*p). However, as more patients who were previously treated with PARPi/platinum therapy are studied in the context of resistance, new gene reversions are being identified. We recently reported a reversion in nibrin (*NBN*) in tumor tissue from a pancreatic acinar cell carcinoma enrolled on TRESR^26^. We evaluated the data from the entire cohort for evidence of reversions in non-HR genes and identified reversion alterations in 2 patients whose tumors harbored biallelic somatic *ATM* PVs (1 breast and 1 colorectal cancer (CRC); **Fig. 5 and Table 2**). Two distinct Class I reversions were identified in each patient. In the CRC patient, available matched buffy coat sequencing confirmed both reversions as somatic. Both patients were treated with platinum-based therapies; the colorectal patient received 1 line of FOLFOX regimen for 11.5 months, while the breast patient had 1 prior line of carboplatin for 4 months. ATM IHC analysis in the diagnostic pre-platinum tissue samples from both patients confirmed ATM protein loss (**Fig. 5b**). No post-platinum tissue samples were available to confirm ATM IHC restoration. To our knowledge these are the first reports of reversions in *ATM*.

**Fig. 5:**
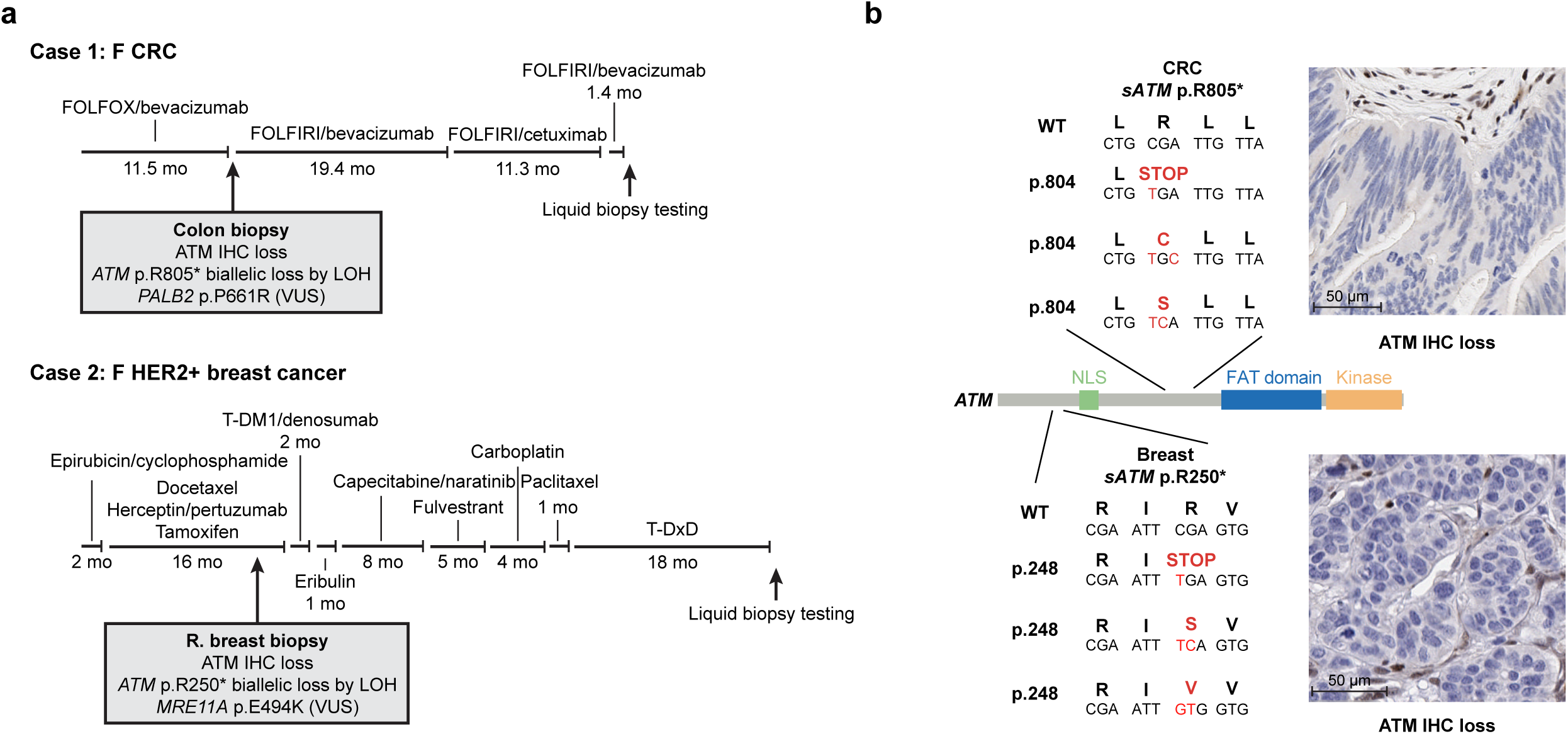
Discovery of *ATM* reversions in ctDNA from two patients. **a)** Prior treatment history and genomic testing for colorectal (top) and breast (bottom) cancer patients with *ATM* reversions. **b)** local sequence changes for the WT, PV and 2 reversions identified in each patient with *ATM* reversion. ctDNA; ctDNA, circulating tumor DNA; CRC, colorectal cancer; F, female; HER2+, human epidermal growth factor receptor 2 positive; IHC, immunohistochemistry; LOH, loss of heterozygosity; mo, month; NLS, nuclear localization signal; PV, pathogenic variant; T-DM1, trastuzumab emtansine; T-DxD, trastuzumab deruxtecan; VUS, variant of unknown significance; WT, wild type.

**Table 2:** Reversion alterations detected in two patients with somatic biallelic ATM alterations.

| Tumor type | Gene | Prior lines | Prior Platinum | Primary |  | Reversion |  |
| --- | --- | --- | --- | --- | --- | --- | --- |
|  |  |  |  | cDNA | AA | Secondary cDNA | Combined AA |
| Breast | ATM | 8 | Carboplatin | c.748C>T | p.R250* | c.749G>C | p.R250S |
|  |  |  |  |  |  | c.748_749delCGins GT | p.R250V |
| CRC | ATM | 4 | 2 lines of FOLFOX | c.2413C>T | p.R805* | c.2415A>C | p.R805C |
|  |  |  |  |  |  | c.2414G>C | p.R805S |

AA, amino acid; cDNA, coding DNA; CRC, colorectal cancer

## DISCUSSION

Herein we present a comprehensive set of ctDNA-based analyses in a pan-tumor cohort of patients with DDR alterations pretreated with PARPi and/or platinum-based chemotherapy. We report the landscape of ctDNA detection of pathogenic variants, methylation-based tumor fraction, ctDNA-based HRD calling and the impact of CH filtering. Using our pipeline, we describe the largest pan-tumor set of mechanisms of resistance to PARPi/platinum therapies and the biological variety and breadth of reversions. We classify these reversions into two groups: Class I, with minimal impact on protein sequence, and Class II, with significant effects on protein sequence and domain structure. A significant proportion of these Class II reversions may have gone undetected by earlier-generation ctDNA-based assays with limited intron coverage of DDR or other putative resistance genes.

We show several key findings related to the utility of ctDNA testing in this context. First, the high confirmation rate reported here for DDR and HR-associated PVs is encouraging; and we extend these data showing that tumor shedding as measured by methylation-based tumor fraction (methyl-TF) is a key factor in the identification of somatic PVs, particularly for LGRs. Second, while variant-based estimates of TF are commonly used, they often overestimate TF due to the small number of variants sampled, and this TF estimate can be greatly impacted by copy number changes or erroneous annotation of CH variants. While most CH variants arise in CH-associated genes, relevant tumor driver genes such as *TP53* and *ATM* are often affected, which can confound the interpretation of ctDNA results^27–30^. The use of methyl-TF dramatically increased the number of samples with quantifiable ctDNA and is not affected by CH-variants suggesting it as the preferred metric for estimating TF. Using methyl-TF, ctDNA was detected in nearly all patients, with higher median TF in prostate compared to ovarian cancer patients, potentially reflecting biological differences in tumor shedding based on sites of frequent metastasis (i.e., bone for prostate cancers vs other organs in peritoneal cavity for ovarian cancers). Third, the ability to detect PV allelic status and genomic signatures such as HRD is an attractive prospect for ctDNA as a diagnostic modality, especially in the context of limited or poor-quality tissue or absence of repeat biopsies at progression. Given the emerging application of these complex biomarkers to ctDNA, additional optimization may be required, and minimal TFs have yet to be established; therefore negative results should be interpreted with caution.

Finally, we identified reversions in *ATM* in two patients previously treated with platinum chemotherapy. Combined with a previous report from the TRESR study from a patient with pancreatic cancer harboring an *NBN* reversion, our data suggest the landscape of reversion alterations extends beyond canonical HRD-associated genes and that reversion analysis should extend to all DDR genes^26^.

The detection rate (44%) of reversion alterations across BRCA-associated tumor types with revertible alterations, along with the high frequency of deletions and LGRs, suggests that the true prevalence of reversions post-PARPi/platinum is higher than previously reported using tumor NGS or earlier generations of ctDNA panels^4–8,11–15^. Reversions have usually been detected at frequencies less than 50% across BRCA-associated tumors and at much lower frequencies in ctDNA where detected reversions are most often SNVs or short deletions^4–8,11–15^.

Commercial ctDNA panels available today typically lack full intronic coverage and often only report deletions and rearrangements for a subset of genes. In this cohort, nearly 30% of reversions detected in ctDNA were LGRs, which was comparable to a previous report using the same technology in analysis of blood from breast cancer patients^13^. In contrast to short deletions, LGRs were largely devoid of microhomology, suggesting the emergence of these LGRs occurred via DNA repair pathways other than MMEJ^31,32^.

While reversions are the most frequently described resistance mechanism, we also identified putative resistance alterations in *TP53BP1* and, to a lesser extent, in *PARP1*, *RIF1*, and *LIG4*. The protein 53BP1, in complex with RIF1, promotes Non-Homologous End Joining (NHEJ) by protecting dsDNA breaks from end-resection^22^. 53BP1 loss is a documented mechanism of PARPi resistance, while mutations in *PARP1* preclude PARP trapping^23^. LIG4 is also directly involved in NHEJ-dependent repair and its loss of function promotes reliance on TMEJ/MMEJ^24,25^. We found that 55% of patients with alternative resistance mechanisms had co-occurring reversions, suggesting that tumors evolve multiple distinct mechanisms of resistance^13^.

This work has several implications for early phase DDR clinical trial design. Multiple DDR-directed therapies are in development, against targets such as ATR, ATM, DNA-PK, DNA polymerase theta, and more, alone or in combination with PARPi, chemotherapy or other agents^2,33^. In most instances, trials for these agents enroll DDR-altered patients who have previously progressed on or are resistant to PARPi or platinum therapies. Our data presented here suggests these are a biologically heterogeneous group, both among and within individual patients. For example, it is conceivable that the benefit of a novel agent may vary depending on whether Class I or Class II reversions are present, with corresponding differences in the degree of restored HR protein function. Similarly, patients without reversions may respond differently to novel agents than patients with reversions. Much of this remains to be delineated in future work, but it is clearly appropriate to abandon the assumption that resistance to PARPi/platinum can be explained by single-gene short-variants that are rarely shared between tumors, even in the same patient. The success and interpretation of future DDR trials may require accurate assessments of HRD and resistance mechanisms from patient samples acquired immediately prior to clinical trial enrollment.

This study has several limitations. First, while modest correlations were observed between baseline methyl-TF and radiographic measurements, non-target lesions were not included in the quantification and therefore the total tumor burden may not accurately be reflected. In prior analysis, when total volume of discrete and non-discrete lesions were considered in a study of patients with HRD-positive metastatic breast cancer, methyl-TF closely mirrored radiological assessment of tumor volume over time^13^. Similarly, the lack of association between baseline TF and CA-125 or PSA is likely driven by interpatient tumor marker variability, which should be studied further. Secondly, tumor shedding and TF vary by tumor type, metastatic burden and treatment history, impacting the detection of reversions, potentially limiting the ability to make accurate comparisons of detection rates. Third, while we observed a high frequency of LGRs and other class II reversions, suggesting that the true prevalence of reversions post-PARPi/platinum is higher than previously reported, we were unable to experimentally validate the functional consequence of these reversions. In this context, recent studies have demonstrated the ability of Class II splice-site reversions to induce exon skipping and restore expression of *BRCA1* and *BRCA2*^13,17^. The functional analysis of HR activity remains experimentally challenging and specific to each patient’s PV and reversion repertoire. Whether Class II reversions fully or partially restore HR activity, remains to be determined. The functional significance of alternative resistance mechanisms reported in this study, while supported by in vitro data^1–3^, have limited clinical validation. Finally, this study does not report on the clinical correlation of the ctDNA covariates described here with outcomes for patients treated with camonsertib given in combination with gemcitabine or PARPi, which will be included in subsequent manuscripts.

This analysis highlights the potential utility of comprehensive genomic and epigenomic ctDNA profiling and the complexity of acquired resistance to standard of care targeted agents in this heterogeneous group of patients. In an era of mechanism-based therapies, future success of DDR therapies will be predicated on the intelligent use of similar data to guide and develop future clinical trials.

## METHODS

### Cohort and sample collection

Between March 25^th^, 2021 and July 17^th^, 2023, blood was collected from subjects enrolled into two Phase 1/2 clinical trials (TRESR; NCT04497116, ATTACC; NCT04972110). Investigational interventions included the ATRi camonsertib alone or in combination with either gemcitabine or PARPi: niraparib, olaparib, or talazoparib. Key eligibility criteria for both trials were the presence of a loss of function genomic alteration in one of seventeen genes predicted to be synthetic lethal with ATR inhibition, including: *ATM*, *ATRIP*, *BRCA1*, *BRCA2*, *CDK12*, *CHTF8*, *FZR1*, *MRE11*, *NBN*, *PALB2*, *RAD17*, RAD50, *RAD51B/C/D*, *REV3L*, *RNASEH2A*, *RNASEH2B*, *SETD2* or other genes agreed upon between the sponsor and investigator as detected by a validated local tissue, plasma or germline test.

The study was conducted in accordance with the Declaration of Helsinki and Council for International Organizations of Medical Sciences International Ethical Guidelines, applicable International Conference on Harmonization, Good Clinical Practice Guidelines, and applicable laws and regulations. All patients provided written informed consent to adhere to the clinical protocol and provided serial blood samples and tumor tissues. The protocol was approved by the Institutional Review Board or Ethics Committee at each participating institution (**Supplemental Table 1**).

Blood for retrospective analysis was collected in two 10 mL K2-EDTA tubes pre-treatment and at one timepoint during each 21- or 28-day cycles. Plasma and buffy coats were isolated immediately following phlebotomy by centrifugation, frozen at −80°C, and bio-banked. Plasma and buffy coat aliquots were processed and analyzed in batches at Guardant Health.

### Cell-free and matched normal DNA sequencing

Upon receipt of frozen plasma, cell-free DNA (cfDNA) was extracted and sequenced at Guardant Health within a College of American Pathologist (CAP) and Clinical Laboratory Approvement Amendments (CLIA) accredited laboratory using the Guardant Infinity^TM^ platform^34^. Guardant Infinity^TM^ was developed, and performance characteristics determined, by the Guardant Health Clinical Laboratory in Redwood City, CA, USA, which is certified under the Clinical Laboratory Improvement Amendments of 1988 (CLIA) as qualified to perform high complexity clinical testing. Guardant Infinity^TM^ here refers to the Research Use Only (RUO) assay. This assay has not been cleared or approved by the US FDA. For the purposes of this study, genomic DNA isolated from buffy coats were sequenced in parallel.

### Methylation-based ctDNA detection and tumor fraction quantification

Methylation-based ctDNA detection was determined using a logistic regression-based model trained on samples from multiple tumor types and cancer-free samples. Tumor fraction quantification was determined using a similar methodology as previously described^13^ but with a model trained on samples from additional tumor types in addition to breast, lung, colorectal, and bladder, and with differentially methylated regions (DMRs) determined from this updated model.

### Variant-based tumor fraction estimation

mVAF and CH-mVAF were used as additional proxies for detection of ctDNA and quantification of TF for comparison to methyl-TF. Detection and quantification of variants were performed as described previously^13,35^. For the purposes of this study, mVAF was defined as the mean VAF among all predicted somatic variants (excluding germline and predicted CH variants, as well as variants that did not have VAF > 0.5% at any timepoint) detected at any timepoint for an individual patient using the Guardant Infinity^TM^ assay. CH-mVAF calculations additionally excluded variants detected in buffy coat samples as well as plasma. mVAF and CH-mVAF calculations were limited to the subset of 74 genes covered by the separate Guardant360^TM^ CDx clinical test^35^. This approach is the basis of genomic-based measures of ctDNA change early after treatment initiation to be described in subsequent manuscripts.

### CH-prediction and CH-detection

For CH detection from paired plasma and buffy coat, a bioinformatics pipeline was used to predict tumor or CH origin of non-germline variants by comparison of molecular support in each of the analytes. Variants with higher allele frequency in buffy coat and variants without statistically significant enrichment in plasma were predicted to be of CH origin, while all other variants detected in plasma were predicted to be of tumor origin. For CH prediction in plasma, a boosted tree model was trained on over 200 plasma samples from cancer-free individuals (N=37) or patients with different types of cancer (N=164). The model was validated and tested on plasma samples from N=129 and N=148 patients with cancer, respectively. The variant labels were obtained from paired buffy coat samples, and the classifier was optimized for high specificity. The model leveraged genomic, methylation, and fragmentomics features, and metadata from over 250,000 historical Guardant Health samples as well as data from COSMIC^36^.

### LOH, cHRD and allelic status determination in plasma

LOH and homozygous deletion calling in plasma was determined using genomic coverage quantification and segmentation^37^, followed by a likelihood-based copy number variation (CNV) caller based on tumor purity and ploidy^38–40^, as previously described^41^. Homozygous deletion calls were defined as loss of all copies of a gene, and LOH calls were defined as copy number loss events with only one allele remaining. HR status and score in plasma were predicted based on a probabilistic logistic regression genomic model that takes into account LOH, large scale transitions, telomeric allelic imbalance, and a single base substitution (SBS)-based signature derived from somatic SNVs detected in breast and prostate cancer HRD samples^42^. An alternate model excluding the SBS signature component is used for samples with less than 10 somatic SNVs. The final ensemble model was trained on breast and prostate cancer samples to predict BRCA1 or BRCA2 deficiency. The HR status is determined by applying a threshold to the probabilistic score which was set to achieve 95% pan-cancer specificity in samples without any genomic alterations in homologous recombination repair (HRR) genes. Enrollment PVs were considered to have detected biallelic LoF if one of the following criteria was met: (1) homozygous deletion; (2) compound heterozygous mutation; (3) mutation and LOH; (4) LGR and LOH; or (5) mutation and non-reversion LGR. Variants predicted to be CH-derived or detected in matched buffy coats were not considered for allelic status determination. Samples without the enrollment PV detected were not considered for allelic assessment.

### Reversion detection

Reversions were identified by the Guardant Health automated pipeline and by manual review. Reversion mutations in HRD-associated genes were defined and detected as previously described^11,43,44^. For manual review, each patient’s Guardant Infinity™ results were reviewed and curated by an expert reviewer. All variants in the enrollment gene were manually classified for the ability to revert mutation sequence using the following criteria:

Secondary variants were determined to be Class I reversions if they met any of the following:

1. Overlapped with the PV and re-established the coding frame with minimal changes to the coding sequence
2. Were non-overlapping but re-established the coding frame with minimal changes to the coding sequence

Secondary variants were classified as Class II reversions if they met any of the following:

1. Abolished a splice-site in or proximal to the PV containing exon resulting in alternative, in-frame splicing
2. Excised exons containing the PV, and the adjacent exons were in-frame
3. Adjusted the frame of an out-of-frame splicing event leading to productive, alternative, in-frame splicing

### Exon splicing enhance/silencer analysis

Intra-exon deletions were queried with Alamut Visual Plus v1.6.1 (Sophia Genetics). The EX-SKIP function was utilized to compare mutant and WT alleles to determine the likelihood of exon skipping^45^.

### Tumor NGS

Archival or contemporaneous tumor Formalin-fixed paraffin-embedded (FFPE) samples were retrospectively sequenced using SNiPDx™, a novel targeted sequencing panel capable of distinguishing monoallelic and biallelic LoF alterations in select DDR genes as previously described^46,47^. DNA was analyzed on a custom anchored multiplex PCR (AMP) panel comprising 26 genes. Amplicon sequencing was performed on the NovaSeq platform (Illumina, San Diego, CA, USA) according to the manufacturer’s standard protocol. Paired-end sequence data were processed using methods developed for AMP to align error-corrected reads^48^. AMP libraries were processed using the VariantPlex Pipeline from Archer Analysis Platform v6.2.8.Archival or contemporaneous tumor FFPE samples were retrospectively sequenced using SNiPDx™, a novel targeted sequencing panel capable of distinguishing monoallelic and biallelic LoF alterations in select DDR genes as previously described^46,47^. DNA was analyzed on a custom AMP panel comprising 26 genes. Amplicon sequencing was performed on the NovaSeq platform (Illumina, San Diego, CA, USA) according to the manufacturer’s standard protocol. Paired-end sequence data were processed using methods developed for AMP to align error-corrected reads^48^. AMP libraries were processed using the VariantPlex Pipeline from Archer Analysis Platform v6.2.8.

Genome-wide major and minor copy numbers were inferred by FACETS^49^. Briefly, copy number alterations and allelic imbalances in the 26 SNiPDx™ target genes and other genomic regions were calculated on the basis of the Log_2_R (i.e., the Log_2_ ratio of single nucleotide polymorphism coverage in a tumor specimen to coverage in a matched panels of normal), and Log_2_ odds ratio (Log_2_OR; calculated from the number of reads reporting the alternative allele:number of reads reporting the reference allele), adjusted by tumor purity and ploidy. Genome-wide major and minor copy numbers were inferred by FACETS^49^.

### Tumor allelic status determination

Tissue-based enrollment PV allelic status was determined by SNiPDx, WGS or local NGS (where available). Enrollment genes were considered to have biallelic LoF if one of the following criteria was met: (1) homozygous deletion; (2) compound heterozygous mutation; (3) mutation and LOH; or (4) mutation and non-overlapping loss. Enrollment genes were considered to have monoallelic loss if the following criteria were met: (1) mutation without LOH or (2) heterozygous loss, considered to have no loss if no mutation or copy number loss was detected. Subclonal alterations were those where the enrollment alteration was detectable at lower-than-expected VAF, upon adjustment for tumor purity, ploidy and local copy number, or present only in some tumor biopsies. If central results were not available and local testing could detect any of the above events leading to biallelic loss, the gene was considered to have biallelic loss. If no result could be determined the allelic status was labeled as unknown.

### Tumor HRD scoring

Tumor HRD scores were derived using allele specific copy number data generated from SNiPDx panel using ovaHRDscar^50^. Briefly, the total number of LOH events between 10-50Mb, large scale transitions (LST) greater than 12Mb and separated by no more than 1Mb, and telomeric allelic imbalances (TAI) that extend up to the telomere but do not cross the centromere, were calculated. No HRD score threshold was applied to tumor NGS data because ovaHRDscar has only been validated on ovarian cancer samples using whole exome sequencing and the empirical threshold (≥54) may not apply to other tumor types or sequencing modalities such as SNiPDx. Tumor sam^≥^ples with a computationally predicted purity of >30% were included in this analysis.

## DATA AVAILABILITY

To minimize the risk of patient re-identification, data will only be shared upon reasonable request. For eligible studies, qualified researchers may request access to individual patient-level clinical data by contacting Repare Therapeutics, Inc. Once approved, the data will be shared through a secure data sharing link.

## CODE AVAILABILITY

Guardant Health ctDNA analysis methods are proprietary. No additional custom algorithms were used for the analysis. Figure generation was performed using R (version 4.1) and tidyverse (version 2.0.0.0.9).

## ACKNOWLEDGMENTS

This work was funded by Repare Therapeutics, Inc., and Guardant Health Inc. Thank you to the patients and families for their participation in clinical research and to Parham Nejad, Ayat Alsaraby and Deepa Parthasarathy (Repare Therapeutics) for ensuring quality biobanking and data acquisition. We thank Adrienne Johnson (Repare Therapeutics) for implementing the ovaHRDScar pipeline and Brooke Overstreet (Guardant Health) for critical manuscript review. Figures created with support from BioRender.com. Figure preparation and submission support were provided by Allison Alwan TerBush, PhD, and Rosie Henderson, MSc, of Onyx (a division of Prime, London, UK).

## AUTHOR CONTRIBUTIONS

**Conception and design:** Ian M. Silverman, Joseph D. Schonhoft, Benjamin Herzberg, Arielle Yablonovitch, Errin Lagow, Sunantha Sethuraman, Michael Zinda, Ezra Y. Rosen, Victoria Rimkunas

**Provision of study materials or patients:** Benjamin Herzberg, Michael Cecchini, Elizabeth Lee, Stephanie Lheureux, Elisa Fontana, Benedito A. Carneiro, Timothy A. Yap, Ezra Y. Rosen

**Collection and assembly of data:** Ian M. Silverman, Joseph D. Schonhoft, Arielle Yablonovitch, Errin Lagow, Sunantha Sethuraman, Danielle Ulanet, Julia Yang, Insil Kim, Paul Basciano, Victoria Rimkunas

**Data analysis and interpretation:** Ian M. Silverman, Joseph D. Schonhoft, Benjamin Herzberg, Arielle Yablonovitch, Errin Lagow, Sunantha Sethuraman, Michael Zinda, Ezra Y. Rosen, Victoria Rimkunas

**Manuscript writing:** All authors

**Final approval of manuscript:** All authors

**Accountable for all aspects of the work:** All authors

## COMPETING INTERESTS

**IMS, JDS, SS, DU, JY, IK, PB, MZ, and VR** are employees of Repare Therapeutics and may hold stock and/or stock options.

Benjamin Herzberg is a study investigator and has received honoraria from Eisai; worked in a consulting or advisory role with Amgen; and received research funding from Repare Therapeutics (Inst), IDEAYA Biosciences (Inst), Amgen (Inst), Revolution Medicines (Inst), and Astellas Pharma (Inst).

Arielle Yablonovitch and Errin Lagow are employees of Guardant Health and may hold stock and/or stock options.

Michael Cecchini is a study investigator and has received a National Cancer Institute (NCI) Mentored Clinical Scientist Research Career Development Award; personal fees from Bayer Pharmaceuticals, DAVA Oncology, Taiho Pharmaceuticals, Seattle Genetics, MacroGenics, and Daiichi Sankyo; and holds stock options from Parthenon Therapeutics.

Elizabeth Lee is a study investigator and reports consulting funding from Aadi Biosciences and OnCusp Therapeutics and has received research funding paid to their institution from Merck, Seagen, Repare Therapeutics, OnCusp Therapeutics, Profound Bio, KSQ Therapeutics/Roche.

Stephanie Lheureux is a study investigator and reports payment or honoraria for lectures, presentations, speaker’s bureaus, manuscript writing or educational events from AstraZeneca, Eisai/Merck, and GSK; and reports consulting fees from AstraZeneca, Eisai, GSK, Merck, Novocure, and Shattuck Labs.

Elisa Fontana is a study investigator and employee of Hospital Corporation of America (HCA) International and has received honoraria from Repare Therapeutics, CARIS Life Science, Seagen, Sapience, BicycleTx Ltd (conference attendance); Astellas, Pfizer (Advisory Board). She has received funding for her institution from Acerta Pharma, ADC Therapeutics, Amgen, Arcus Biosciences, Array BioPharma, Artios Pharma Ltd, Astellas Pharma Inc, Astex, Astra Zeneca, Basilea, Bayer, BeiGene, BicycleTx Ltd, BioNTech, Blueprint Medicines, Boehringer Ingelheim, Calithera Biosciences, Inc., Carrick Therapeutics, Casi Pharmaceuticals, Clovis Oncology, Inc, Crescendo Biologics Ltd., CytomX Therapeutics, Daiichi Sankyo, Deciphera, Eli Lilly, Ellipses, Exelixis, F. Hoffmann-La Roche Ltd, Fore Biotherapeutics, G1 Therapeutics, Genentech, GSK, H3 Biomedicine Inc, Hutchinson MediPharma, Ignyta/Roche, Immunocore, Immunomedics, Inc., Incyte, Instil Bio, IOVANCE, Janssen, Jiangsu Hengrui, Kronos Bio, Lupin Limited, MacroGenics, Menarini, Merck KGaA, Mereo BioPharma, Merus, Millennium Pharmaceuticals, MSD, Nerviano Medical Sciences, Nurix Therapeautics Inc, Oncologie, Oxford Vacmedix, Pfizer, Plexxikon Inc., QED Therapeutics, Inc., Relay Therapeutics, Repare Therapeutics, Ribon Therapeutics, Roche, Sapience, Seagen, Servier, Stemline, Synthon Biopharmaceuticals, Taiho, Tesaro, Turning Point Therapeutics, Inc, PMVPharma, and Takeda.

Benedito A. Carneiro is a study investigator and has received research funding paid to their institution by Abbvie, Actuate Therapeutics, Agenus, Astellas, AstraZeneca, Bayer, Daiichi Sankyo, Dragonfly Therapeutics, Pfizer Inc., Pyxis Oncology, and Repare Therapeutics.

Jorge S. Reis-Filho has received consultancy fees from Goldman Sachs, Paige.AI, Repare Therapeutics, and Personalis; is a member of the scientific advisory boards of Volition Rx, Paige.AI, Repare Therapeutics, Personalis, and Bain Capital; is a member of the board of directors of Grupo Oncoclinicas; and is an ad hoc member of the scientific advisory boards of Roche Tissue Diagnostics, Ventana Medical Systems, AstraZeneca, Daiichi Sankyo, and Merck Sharp & Dohme.

Timothy A. Yap is a study investigator and an employee of The University of Texas MD Anderson Cancer Center, where he is Vice President, Head of Clinical Development in the Therapeutics Discovery Division, which has a commercial interest in DDR and other inhibitors (IACS30380/ART0380 was licensed to Artios); has received funding paid to their institution from Acrivon, Artios, AstraZeneca, Bayer, BeiGene, BioNTech, Blueprint, Bristol Myers Squibb, Boundless Bio, Clovis, Constellation, Cyteir, Eli Lilly, EMD Serono, Forbius, F-Star, GlaxoSmithKline, Genentech, Haihe, Ideaya ImmuneSensor, Insilico Medicine, Ionis, Ipsen, Jounce, Karyopharm, KSQ, Kyowa, Merck, Mirati, Novartis, Pfizer, Ribon Therapeutics, Regeneron, Repare, Rubius, Sanofi, Scholar Rock, Seattle Genetics, Tango, Tesaro, Vivace, and Zenith; has received consultancy funding from AbbVie, Acrivon, Adagene, Almac, Aduro, Amphista, Artios, Astex, AstraZeneca, Athena, Atrin, Avenzo, Avoro, Axiom, Baptist Health Systems, Bayer, BeiGene, BioCity Pharma, Blueprint, Boxer, Bristol Myers Squibb, C4 Therapeutics, Calithera, Cancer Research UK, Carrick Therapeutics, Circle Pharma, Clovis, Cybrexa, Daiichi Sankyo, Dark Blue Therapeutics, Diffusion, Duke Street Bio, 858 Therapeutics, EcoR1 Capital, Ellipses Pharma, EMD Serono, Entos, F-Star, Genesis Therapeutics, Genmab, Glenmark, GLG, Globe Life Sciences, GSK, Guidepoint, Ideaya Biosciences, Idience, Ignyta, I-Mab, ImmuneSensor, Impact Therapeutics, Institut Gustave Roussy, Intellisphere, Jansen, Kyn, MEI pharma, Mereo, Merck, Merit, Monte Rosa Therapeutics, Natera, Nested Therapeutics, Nexys, Nimbus, Novocure, Odyssey, OHSU, OncoSec, Ono Pharma, Onxeo, PanAngium Therapeutics, Pegascy, PER, Pfizer, Piper-Sandler, Pliant Therapeutics, Prolynx, Radiopharma Theranostics, Repare, resTORbio, Roche, Ryvu Therapeutics, SAKK, Sanofi, Schrodinger, Servier, Synnovation, Synthis Therapeutics, Tango, TCG Crossover, TD2, Terremoto Biosciences, Tessellate Bio, Theragnostics, Terns Pharmaceuticals, Tolremo, Tome, Thryv Therapeutics, Trevarx Biomedical, Varian, Veeva, Versant, Vibliome, Voronoi Inc, Xinthera, Zai Labs, and ZielBio; and is a stockholder in Seagen. He was supported by the NCI Cancer Center Support Grant CA016672 to The University of Texas MD Anderson Cancer Center, DOD grants W81XWH2210504_BC211174 and W81XWH-21-1-0282_OC200482, V Foundation Scholar Grant VC2020-001, and NIH R01 grant 1R01CA255074.

Ezra Y. Rosen is a study investigator.

