## Supplementary Figures for "Comprehensive baseline ctDNA characterization in two biomarker-selected Phase 1/2 studies using genomic and methylation profiling"

**Supplemental Table and Figures for:**

**Supplemental Table 1. Institutional Review Boards of participating institutions**

| **TRESR (NCT04497116)** | |  |
| --- | --- | --- |
| **IRB Name** | **Affiliation(s)** | **Country** |
| University of Texas MD Anderson Cancer Center Institutional Review Boards | MD Anderson Cancer Center | USA |
| Dana Farber Cancer Institute Institutional Review Boards | Dana-Farber Cancer Center | USA |
| Advarra IRB, Inc. | Sarah Cannon Research Institute - TN | USA |
| Memorial Sloan-Kettering Cancer Center Institutional Review Board/Privacy Board | Memorial Sloan Kettering Cancer Center | USA |
| Duke University Health Systems Institutional Review Boards | Duke Cancer Institute | USA |
| Rhode Island Hospital IRB | Rhode Island Hospital | USA |
| Northwestern University IRB Panels A, B, C, D and Q | Robert H. Lurie Comprehensive Cancer Center of Northwestern University | USA |
| University Health Network Research Ethics Board | Princess Margaret Cancer Centre | Canada |
| Health Research Authority, North East – Tyne and Wear South Research Ethics Committee | The Christie NHS Foundation Trust Manchester  Freeman Hospital Newcastle/Sir Bobby Robson Cancer Trials Research Centre  Sarah Cannon Research Institute – London | United Kingdom |
| Scientific Ethics Committees for the Capital Region (Denmark) – *translated name* | Rigshospitalet, University Hospital of Copenhagen | Denmark |
| **ATTACC (NCT04972110)** | | |
| **IRB name** | **Affiliation(s)** | **Country** |
| Columbia University Medical Center Institutional Review Board | Columbia Herbert Irving Comprhensive CA Ctr (CUHC) | USA |
| Johns Hopkins Institutional Review Board | Johns Hopkins University (JHU) | USA |
| Advarra IRB, Inc. | Yale Cancer Center  Huntsman Cancer Institute and Hospital  University of Michigan Rogel Cancer Center  Thomas Jefferson University | USA |
| Mayo Clinic Institutional Review Board | Mayo Clinic - Rochester  Mayo Clinic – Arizona  Mayo Clinic - Florida | USA |
| UCSF Human Research Protection Program & IRB | University of California, San Francisco Helen Diller Family Comprehensive Cancer Center | USA |
| US Oncology Inc., IRB | Rocky Mountain Cancer Centers, LLP  Oncology Associates of Oregon, P.C | USA |

**Comprehensive baseline ctDNA characterization in two biomarker-selected Phase 1/2 studies using genomic and methylation profiling**

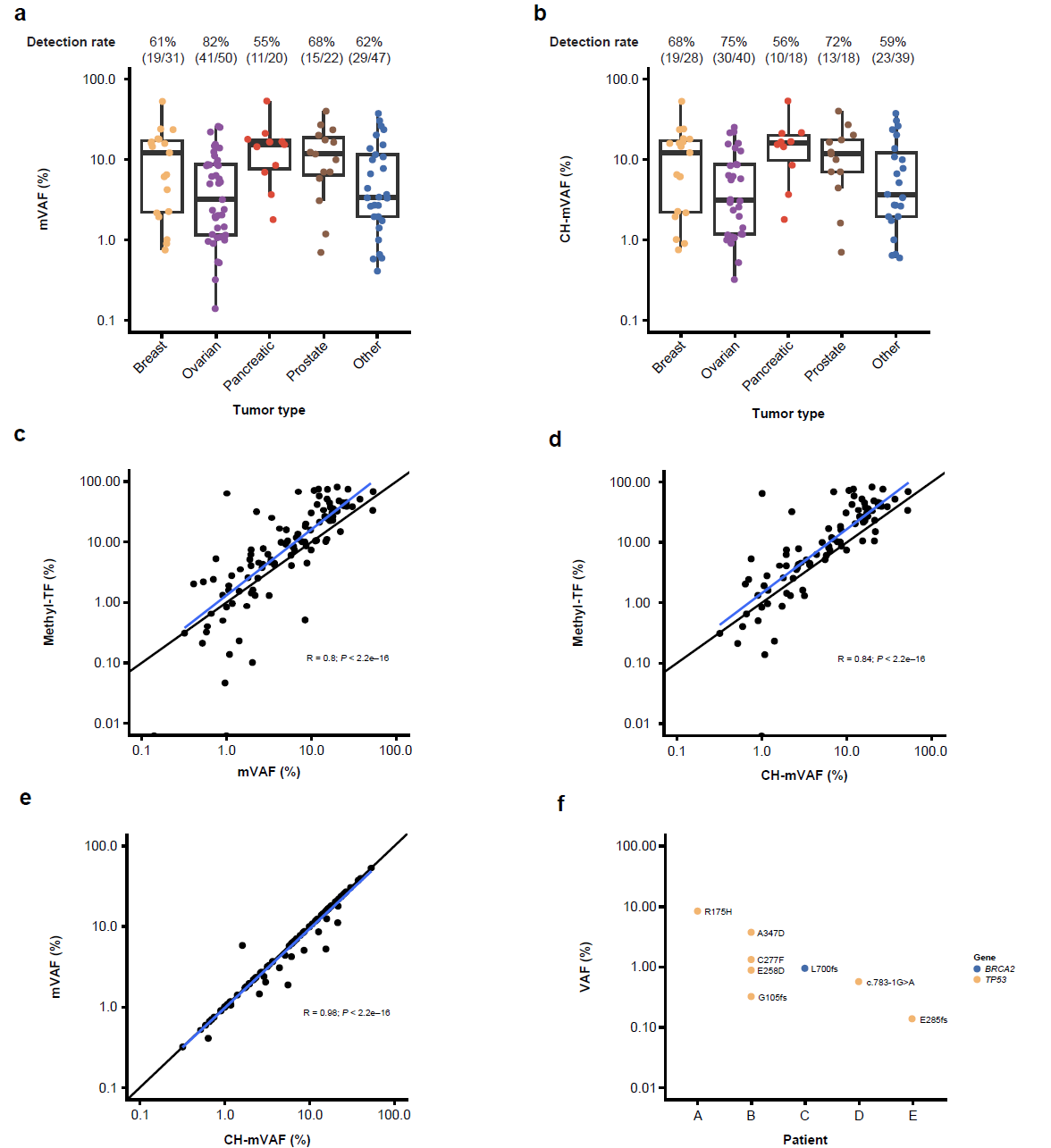

### **Supplemental Fig. 1. Relationship between genomic and epigenomic measures of tumor fraction.**

**a, b)** Detectability and range of ctDNA, mVAF, and CH-filtered mVAF utilizing the Guardant Infinity™ assay and focused on the subset of 74–genes covered by the current Guardant360^®^ CDx test. **c, e)** Correlations between methyl-TF, mVAF, and CH-mVAF. The black line denotes the diagonal, while the blue lines represent the best-fit linear line. Pearson correlation coefficients are shown. **f)** In samples from 5 patients variants used to calculate mVAF were detected exclusively in the buffy coat.

CH, clonal hematopoiesis; ctDNA, circulating tumor DNA; methyl-TF, methylation-based TF; mVAF, mean variant allele frequency; TF, tumor fraction.

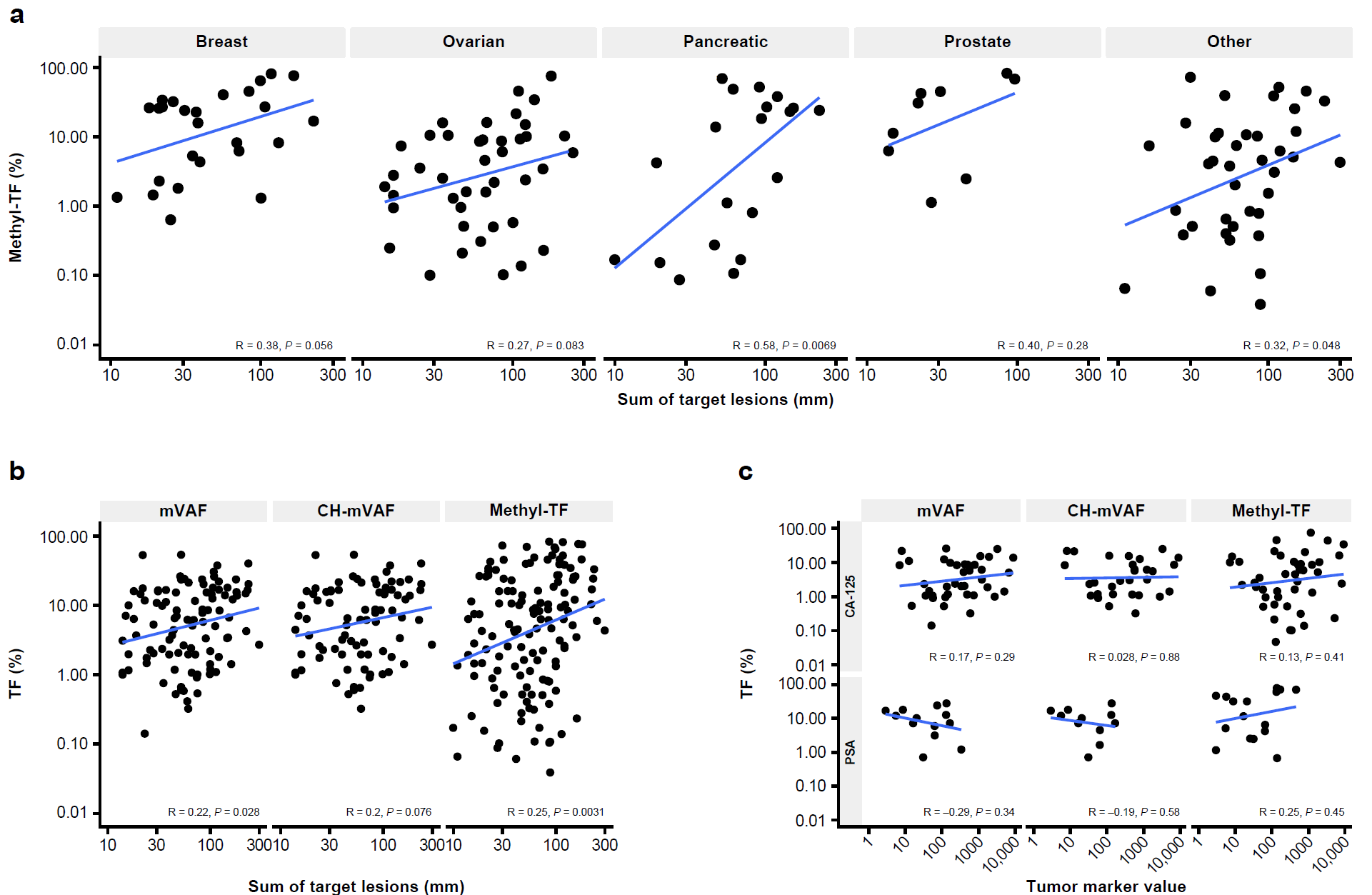

### **Supplemental Fig. 2. Relationship between TF measures and clinical disease burden.**

**a)** Correlation between methyl-TF and baseline sum of target lesion diameters by tumor type. **b, c)** Correlation between baseline TF estimates (mVAF, CH-mVAF and methyl-TF) and **b**) Sum of target lesion diameters and **c**) Baseline CA-125 (ovarian cancer) and PSA (prostate cancer) tumor marker values.

CA-125, cancer antigen 125; CH, clonal hematopoiesis; methyl-TF, methylation-based TF; mVAF, mean variant allele frequency; PSA, prostate-specific antigen; TF, tumor fraction

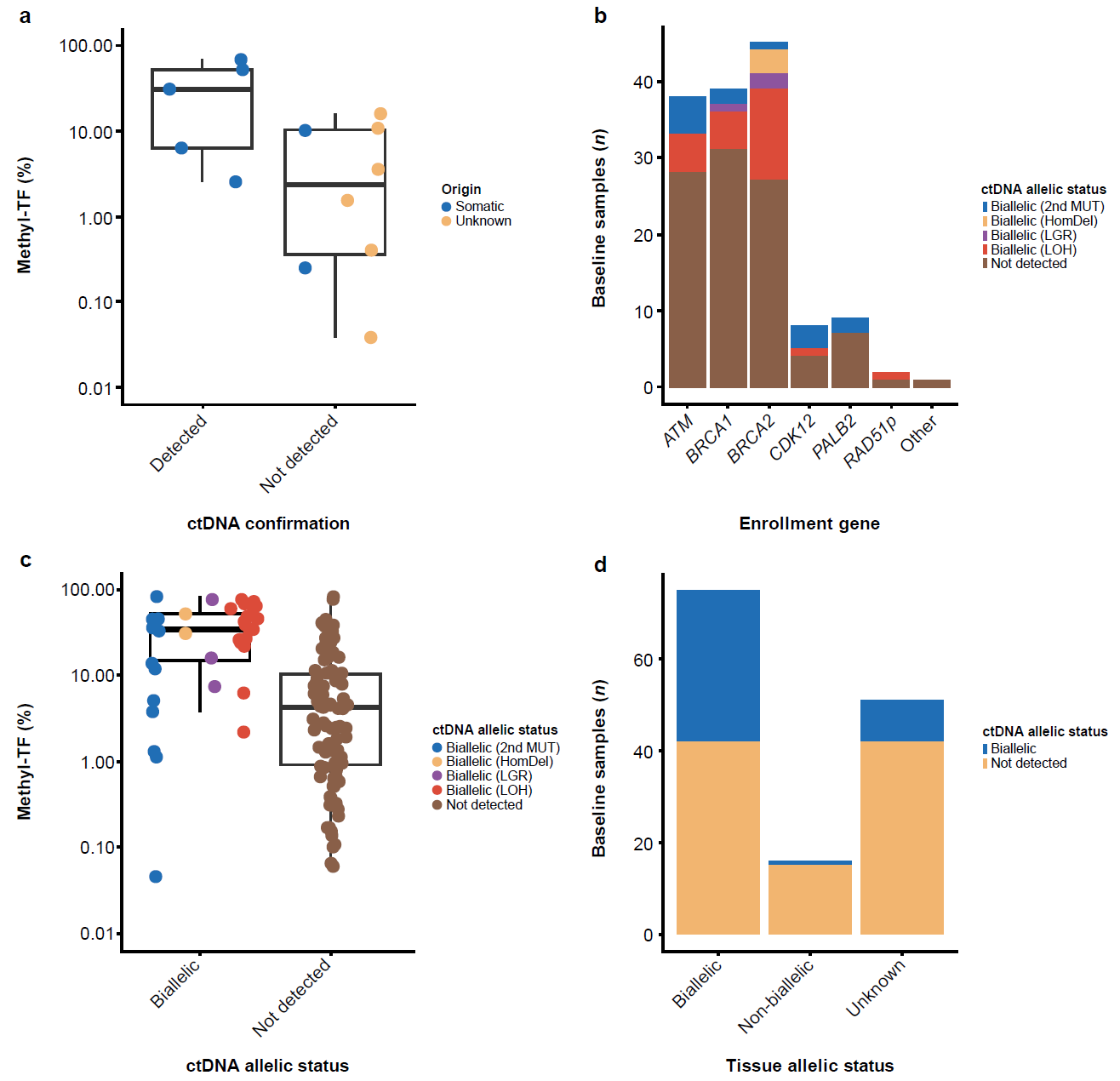

### **Supplemental Fig. 3. ctDNA-based enrollment PV detection and allelic status calls.**

**a)** methyl-TF in patients with detected vs. not detected LGR enrollment PVs of somatic or unknown origin. **b)** Frequency of ctDNA based biallelic status by enrollment PV gene. **c)** methyl-TF values in patients with confirmed biallelic status in ctDNA versus those that biallelic status was not able to be determined. **d)** Relationship between tissue-based and ctDNA-based allelic status.

ctDNA, circulating tumor DNA; HomDel, homozygous deletion; LGR, large genomic rearrangements; LOH, loss of heterozygosity; methyl-TF; methylation-based TF; MUT, mutant; PV, pathogenic variant; TF, tumor fraction.

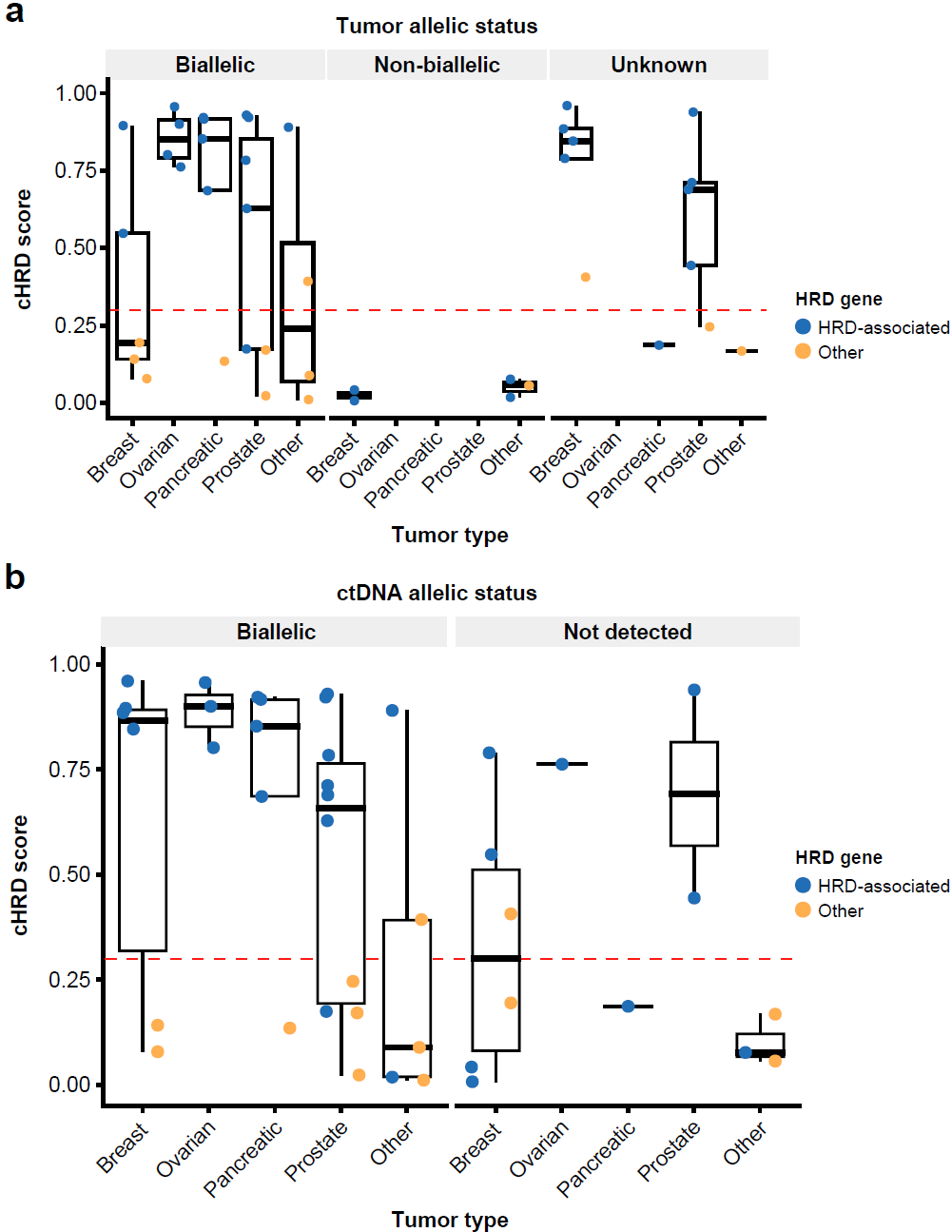

**Supplemental Fig. 4. cHRD scores based on tumor type, genotype and tumor or ctDNA-derived allelic status.**

**a, b)** cHRD scores vs. methyl-TF by enrollment gene, BRCA-associated tumor class and **a)** Tumor-derived or **b)** ctDNA-derived allelic status for samples with >25% methyl-TF. cHRD positivity threshold is 0.3 (horizontal red line).

cHRD, ctDNA-based HRD; ctDNA, circulating tumor DNA; HRD, homologous recombination deficiency; methyl-TF, methylation-based TF; TF, tumor fraction; vs., versus.

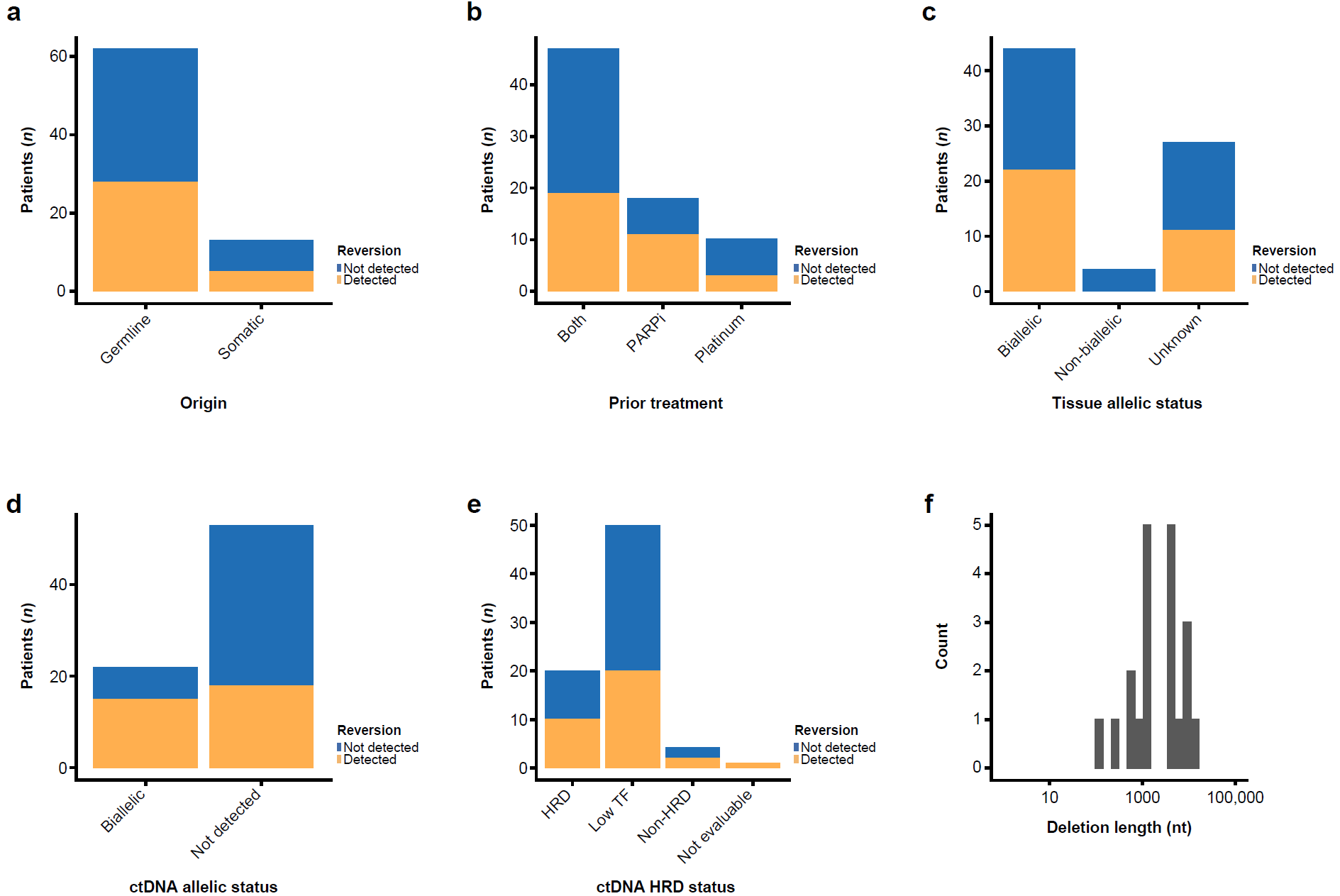

### **Supplementary Fig. 5. Reversion detection by clinical and genomic characteristics.**

**a-e)** Reversion detection by **a)** PV origin, **b)** Prior PARPi/Platinum therapy, **c)** Tissue-derived allelic status, **d)** ctDNA-derived allelic status and **e)** cHRD status. **f)** Distribution of LGR deletion reversion length

cHRD, ctDNA-based HRD; ctDNA, circulating tumor DNA; HRD, homologous recombination deficiency; LGR, large genomic rearrangements; nt, nucleotide; PARPi, Poly (ADP-ribose) polymerase inhibitor; PV, pathogenic variant; TF, tumor fraction.
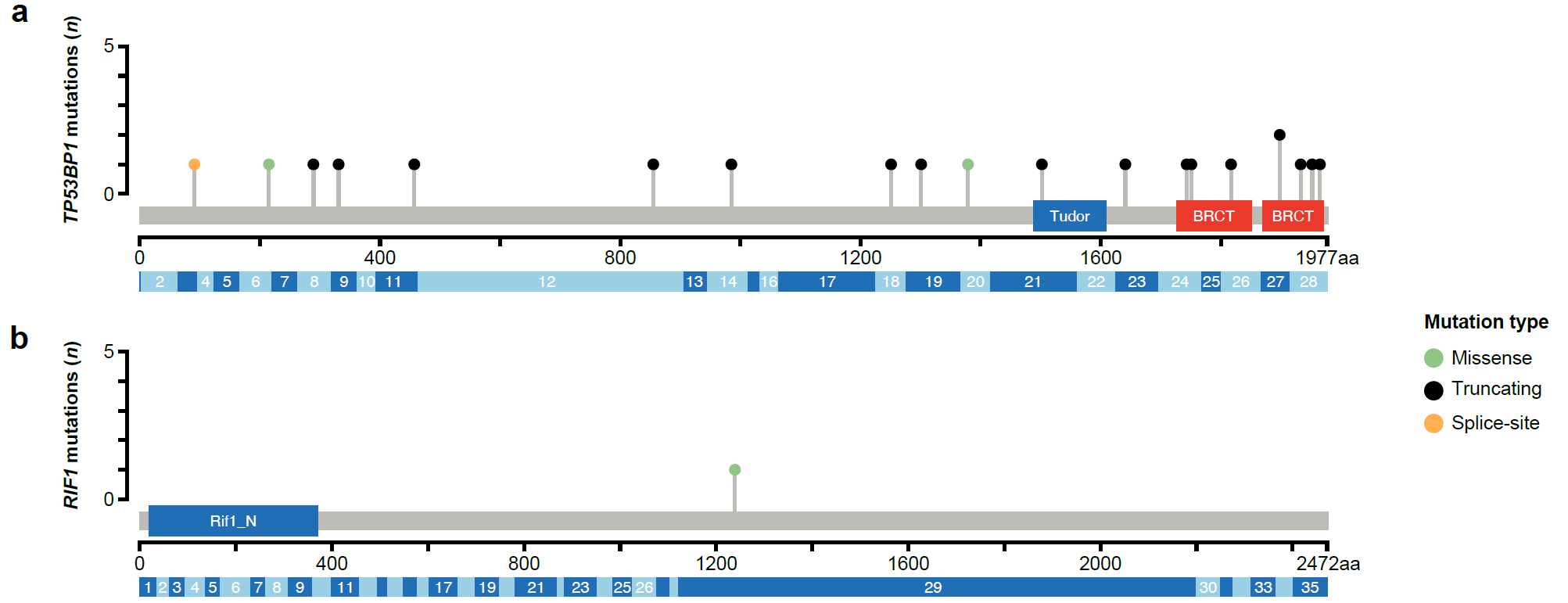

### **Supplementary Fig. 6. Polyclonal *TP53BP1* mutations in a patient with *BRCA1* pancreatic cancer.**

**a, b)** Lollipop plot of the **a)** *TP53BP1* and **b)** *RIF1* mutations identified in a patient with *BRCA1* altered pancreatic cancer. Green indicates missense, black indicates truncating and orange indicates splice-site mutation.

aa, amino acid; BRCT, BRCA1 c-terminal; Rif1_N, Rap1-interacting factor 1 N terminal.
